# Impact of the early stages of the COVID-19 pandemic on coverage of RMNH interventions in Ethiopia

**DOI:** 10.1101/2021.08.24.21262554

**Authors:** Emily Carter, Linnea Zimmerman, Ellie Qian, Tim Roberton, Assefa Seme, Solomon Shiferaw

## Abstract

**Background:** The COVID-19 pandemic and response have the potential to disrupt access and use of reproductive, maternal, and newborn health (RMNH) services. Numerous initiatives aim to gauge the indirect impact of COVID-19 on RMNH.

**Methods:** We assessed the impact of COVID-19 on RMNH coverage in the early stages of the pandemic using panel survey data from PMA-Ethiopia. Enrolled pregnant women were surveyed 6-weeks post-birth. We compared the odds of service receipt, coverage of RMNCH service indicators, and health outcomes within the cohort of women who gave birth prior to the pandemic and the COVID-19 affected cohort. We calculated impacts nationally and by urbanicity.

**Results:** This dataset shows little disruption of RMNH services in Ethiopia in the initial months of the pandemic. There were no significant reductions in women seeking health services or the content of services they received for either preventative or curative interventions. In rural areas, a greater proportion of women in the COVID-19 affected cohort sought care for peripartum complications, ANC, PNC, and care for sick newborns. Significant reductions in coverage of BCG vaccination and chlorohexidine use in urban areas were observed in the COVID-19 affected cohort. An increased proportion of women in Addis Ababa reported postpartum family planning in the COVID-19 affected cohort. Despite the lack of evidence of reduced health services, the data suggest increased stillbirths in the COVID-19 affected cohort.

**Discussion:** The government of Ethiopia’s response to control the COVID-19 pandemic and ensure continuity of essential health services appears to have successfully averted most negative impacts on maternal and neonatal care. This analysis cannot address the later effects of the pandemic and may not capture more acute or geographically isolated reductions in coverage. Continued efforts are needed to ensure that essential health services are maintained and even strengthened to prevent indirect loss of life.

**What is already known?:**

- COVID-19 pandemic and response have the potential to disrupt access and use of reproductive, maternal, and neonatal health services
- Anecdotal evidence suggests some disruptions to health system staffing and resources, service access, and health campaigns in Ethiopia early in the pandemic

**What are the new findings?:**

- Our analysis of PMA-Ethiopia panel survey data shows little disruption of RMNH services in Ethiopia in the initial months of the pandemic
- Compared to immediately prior to the pandemic we observed an increase in care-seeking in rural areas, commodity-related intervention reductions in urban areas, and an increase in postpartum family planning in Addis Ababa
- Despite the lack of evidence of a reduction in health services, the data suggest increased stillbirths in the COVID-19 affected cohort

**What do the new findings imply?:**

- The government of Ethiopia successfully maintained continuity of most RMNCH services during the early stages of the COVID-19 pandemic
- Continued efforts are needed to ensure that essential health services are maintained and even strengthened to prevent indirect loss of life

## Background

The first case of COVID-19 in Ethiopia was reported on March 13, 2020 (1). Followed by early preventative measures such as mandatory quarantine for travelers, mask mandates, and communication efforts, the government of Ethiopia declared a national state of emergency on April 8, 2020 (2). The Ethiopian Federal Ministry of Health swiftly put in place a series of national COVID-19 response policies, notably, with a focus to maintain essential health services, including reproductive, maternal, and newborn health (RMNH) services (3). However, despite national guidelines to sustain essential health services during the pandemic, it is uncertain to what extent these guidelines were adopted by state and local authorities. Other supply and demand-side challenges also complicated the potential impact of COVID-19 on RMNH intervention coverage in Ethiopia.

On the supply side, many health facilities needed to re-allocate medical resources and personnel to emergency responses, potentially leading to a reduction in the availability and quality of non-COVID services (4–6). Staff shortage and nosocomial COVID-19 infection likely created burnout in the health workforce (7). Additionally, following the declaration of a national emergency, Ethiopia postponed nationwide routine vaccination campaigns and scheduled supplemental immunization activities (8). Other RMNH services delivered through campaigns were likely similarly disrupted.

Governmental restrictions on movement and limited access to transportation created barriers in accessing RMNH services on the demand side. There is evidence that these challenges disproportionately affected the most vulnerable groups who lived on daily wages (9). Although health facilities remained open during the pandemic, there is evidence of a decline in utilization of services in public hospitals due to fear of COVID-19 infection (4,9,10).

The potential supply and demand side challenges potentially contribute to disruption in RMNH services, that put the health and wellbeing of mothers and children at risk. It has been estimated that even the most conservative prediction of RMNH service coverage reduction would lead to 253,500 additional child deaths and 12,200 additional maternal deaths over 6 months in low- and middle-income countries (11).

Recognizing the urgency of maintaining RMNH services amid the pandemic, several global efforts have focused on ensuring service coverage and monitoring disruption in services. For example, the WHO “pulse” survey documented widespread disruptions in essential health services across the globe, with greater disruptions reported in low- and middle-income countries. Routine immunization, family planning, and antenatal care services were among the most frequently disrupted services (12). While providing valuable overviews of trends in service availability since COVID-19, this tool is subject to self-report and selection bias and does not reflect the lived experiences of those impacted by the pandemic. An alternative approach to monitoring changes in health service coverage during COVID-19 is to use routine data from health management information systems (HMIS) (13). In theory, HMIS provides “real-time” tracking of the coverage and quality of a range of health services. However, persistent challenges related to lags in reporting, poor/inconsistent data quality, and incomplete data due to the pandemic limit HMIS data’s ability to provide reliable estimates (13–15). Lastly, the World Bank supported several efforts to use phone-based surveys to assess the impacts of COVID-19 on households and individuals. Results from this high-frequency monitoring confirmed the reduction in care-seeking due to fear of COVID-19 exposure or stay-at-home orders. However, this tool also faces challenges related to non- response, under-coverage of vulnerable population, and limited capacity to collect detailed responses (16).

The objective of the current study is to assess changes in intervention coverage in the peripartum period using the Performance Monitoring for Action Ethiopia longitudinal data. By comparing RMNH service utilization and birth outcomes between a COVID-19 unaffected cohort with those potentially impacted by COVID-19, this study provides insights on the effect of the COVID-19 on essential RMNH intervention coverage in Ethiopia.

## Methods

### Data source

Data for this study come from the Performance Monitoring for Action (PMA) Ethiopia survey, a survey project comprised of an annual nationally representative cross-sectional survey, a panel survey following women from pregnancy through one year, and an annual Service Delivery Point (SDP) surveys. The data for this analysis come from the panel survey. PMA Ethiopia is conducted in collaboration between Addis Ababa University and Johns Hopkins Bloomberg School of Public Health.

PMA Ethiopia panel survey used a multistage cluster sampling using probability proportional to size to select 217 enumeration areas (EAs) across six regions in Ethiopia, with region (Afar, Addis Ababa, Amhara, Oromia, Southern Nations Nationalities and Peoples, and Tigray) and residence (urban/rural) as strata. In Afar and Addis Ababa, only region was used for strata. To identify women for the panel survey, a census was conducted among 36,614 households between October and November 2019. All women aged 15-49 were screened (32,792) and, if they reported being currently pregnant or having delivered within the past six weeks, were eligible for the panel study; 2,889 women were identified as eligible and 2,855 enrolled to complete interviews at enrollment, six weeks, six months, and one year postpartum (Figure 1). Data used in this paper were reported at the six-week interview, which had a follow-up rate of 93.3%.

**Figure 1.**
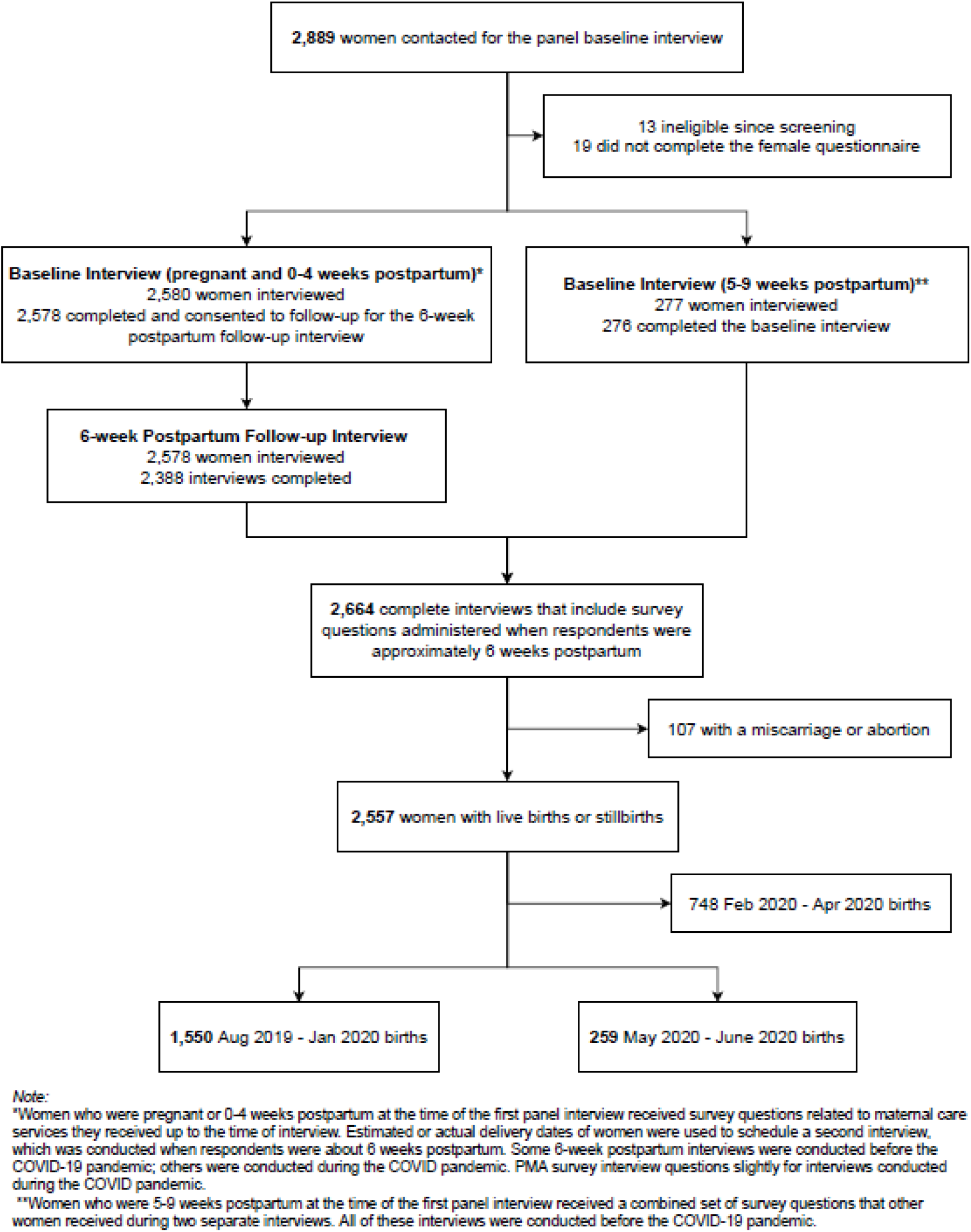
Study Cohort Diagram.

PMA Ethiopia paused data collection in early April due to the COVID-19 pandemic. At that time, questionnaires were modified to include a range of questions about COVID-19 knowledge and risk and the role of COVID-19 in care-seeking behaviors for MNH. When data collection resumed in June with enhanced safety protocols, including social distancing, COVID-19 symptom screening, and mandatory mask requirements, all women with outstanding surveys were interviewed using the updated questionnaires. As a sub-cohort of women had delivered prior to the onset of the COVID-19 pandemic and a sub-cohort delivered during the COVID-19 pandemic, a “natural experiment” within the PMA Ethiopia cohort was introduced, providing a unique opportunity to apply a pre-post cross-sectional study design to examine the early impact of COVID-19 on the coverage of peripartum care indicators.

### Ethical approval

Women provided oral consent to participate at the initial household screening and prior to enrollment in the panel survey for all eligible women. All procedures were approved by both the Addis Ababa University [075/13/SPH] and Johns Hopkins Bloomberg School of Public Health [00009391] Institutional Review Boards. Additional information on the PMA Ethiopia survey can be found at Zimmerman 2020 (17).

### Definition of COVID-19 unaffected and affected cohorts

Restrictions to curb the spread of COVID-19 were introduced in Ethiopia between last March and early April, with some variation in date of introduction by regional states. In addition to structural disruptions, we assume this time also aligns with an increased public awareness of the potential threat of COVID-19. Translating this period of restriction into potential impact on health service access and use in the PMA cohort, we assume those women who gave birth in April or later could experience disruptions to late- ANC visits, care offered during childbirth, and services delivered in the first month after birth. If restrictions did impact service availability, we expect it would immediately affect labor and delivery care. Impact on ANC would be tempered due to repeat service visits throughout the pregnancy. For births that occurred in May 2020, disruption to antenatal service would translate to potential loss of the final pre-birth visit under a four-visit ANC schedule. Care delivered in the neonatal period could also have been impacted in births occurring as early as March 2020.

In defining the appropriate COVID-19 affected and unaffected groups, we also considered the comparability of recall periods. Due to a pause in six-week post-birth follow-up interviews in April and May, births between February and April received follow-up interviews up to 25 weeks after birth (Supplemental Fig 1). This delay in follow-up could result in lower recall accuracy across indicators and significant bias in indicators with reference periods tied to the timing of interview administration (e.g., current breastfeeding or family planning use) or time between birth and interview (e.g., care-seeking for illness in newborn since birth). For our primary analysis, we defined our COVID-19 affected cohort as those born in May 2020 (average recall period: 6.8 weeks) or later and our COVID-19 unaffected cohort as births between August 2019 (start of post-birth data collection) and January 2020 (average recall period: 9.4 weeks). Births that occurred between February and April 2020 were excluded.

We conducted a sensitivity analysis of indicators with a time-invariant reference period more loosely defining the unaffected cohort as August 2019 to February 2020 births (average recall period: 8.6 weeks) and the COVID-19 affected cohort as births in April 2020 or later (average recall period: 12.0 weeks). For indicators with unrestricted reference periods, therefor most susceptible to bias due to differences in recall period (i.e., vaccination, exclusive breastfeeding, care-seeking for infant illness, and postpartum family planning), we restricted the comparison of cohorts to only follow-up interviews that occurred more than five weeks and less than ten weeks after birth (mean recall period COVID-19 unaffected cohort: 6.7 weeks; COIVD-19 affected cohort: 7.9 weeks).

### Indicators of care across the MNH continuum of care

We examined the effect of the COVID-19 pandemic and response on health interventions in the peripartum period. The PMA Ethiopia six-week questionnaire collected data on standard indicators of health practices and interventions during antenatal care, childbirth, and the neonatal period. Where an intervention could only be received through contact with the formal health system (e.g., blood transfusion) we report the indicator as the proportion of the population accessing the service that received the intervention. These indicators serve to assess changes in the content (and potentially quality) of service administered during the time period. Indicators of service contact (e.g., facility delivery) are calculated as a proportion of the total target population and demonstrate potential changes in both care-seeking behaviors and service access. Interventions or practices that can be accessed through multiple healthcare channels or do not require engagement with the healthcare system are similarly presented as coverage indicators among the total target population.

### Data analysis

To assess the effect of the COVID-19 pandemic and response on health practices, services, and outcomes, we compared these indicators in our COVID-19 affected cohort versus our unaffected reference cohort. The primary analysis estimated the odds ratio of intervention receipt or practice (yes/no) for those in the COVID-19 affected cohort compared to the reference cohort using logistic regression. We calculated the association at the national level, with and without adjusting for characteristics of the mother and birth. The adjusted regression assessed the cohort effect after accounting for variations in parity (first birth, 1-2 previous births, 3+ births), maternal education (none, attended primary, attended secondary or higher), maternal age, household wealth (relative quintile), urban versus rural residence, and regional state.

We also looked at associations between cohorts residing in Addis Ababa, other urban areas, and rural areas separately, with and without adjusting for covariates. We posited restrictions and COVID burden might have a greater impact in population centers that are more dependent on public transport, more vulnerable to economic shocks, and more susceptible to COVID-19 transmission. We also calculated the unadjusted coverage of each intervention or practice in both the COVID-19 affected and unaffected cohorts.

We also compared the incidence of stillbirth and neonatal death in the two cohorts, using Poisson regression. To account for potential left truncation of our data due to the absence of early stillbirths among women enrolled late in pregnancy, we restricted our stillbirth analysis to only those enrolled in either their first or second trimester of pregnancy.

As a secondary analysis of an existing data set, neither patients nor the public were involved in the design, conduct, reporting, or dissemination plans of our analysis.

## Results

We analyzed data on health interventions collected six weeks after birth for 1809 women, including 1550 women who gave birth between August 2019 and January 2020 (reference cohort) and 259 women who gave birth in May 2020 or later (COVID-19 affected cohort) (Supplemental Fig 2). In the reference cohort, the 1550 pregnancies resulted in 1506 singleton live births, 17 singleton stillbirths, 26 sets of liveborn twins, and twins with one stillborn. Among the women in the COVID cohort, the 259 pregnancies resulted in 13 stillbirths, 243 singleton live births, and three sets of liveborn twins. The cohorts were similar in maternal education and age, household wealth, and regional distribution (Table 1). However, the COVID cohort included a greater proportion of primiparous and rural women.

**Table 1.**
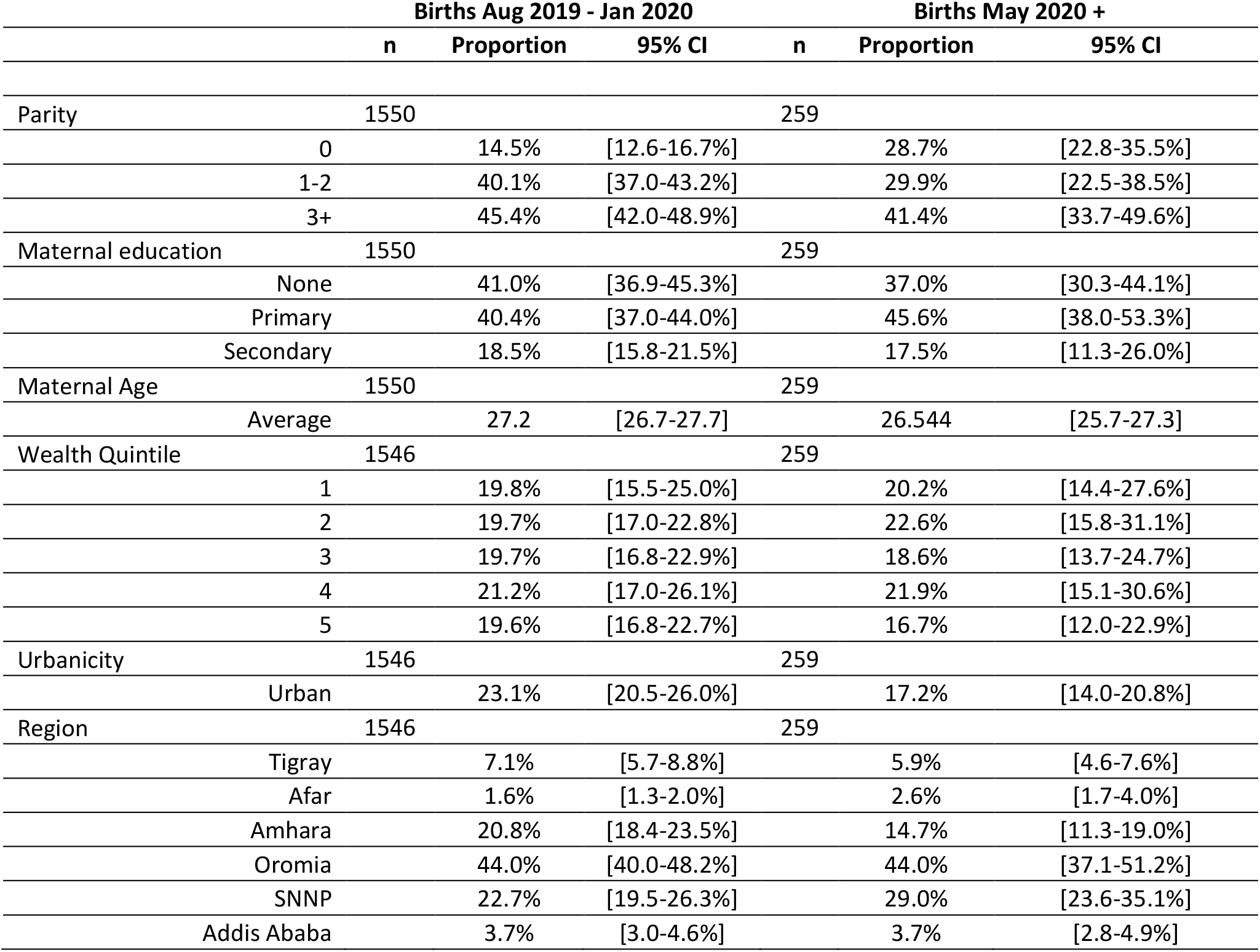
Characteristics of cohorts.

The adjusted and unadjusted odds ratio of intervention receipt or practice in the COVID-19 affected cohort versus the unaffected cohort at the national level is presented in Table 2. Table 3 presents the adjusted and unadjusted odds ratios stratified by urbanicity, including Addis Ababa, other urban areas, and rural areas. Unadjusted estimates of intervention coverage in the COVID-19 affected cohort versus unaffected reference cohort are presented at the national level (Supplemental Table 1) and stratified by residence (Supplemental Table 2).

**Table 2.** Odds of intervention receipt in COVID-19 impacted cohort versus unaffected reference cohort at national level.

|  | Unadjusted |  |  | Adjusted |  |  |
| --- | --- | --- | --- | --- | --- | --- |
|  | n | OR | 95% CI | n | AOR | 95% CI |
| Women with 4+ ANC visits | 1809 | 1.35 | [0.96-1.90] | 1809 | 1.53 | [1.05-2.23] |
| Among women with any ANC: |  |  |  |  |  |  |
| BP check | 1361 | 1.06 | [0.61-1.86] | 1361 | 1.09 | [0.61-1.93] |
| Weighed | 1361 | 1.15 | [0.71-1.89] | 1361 | 1.23 | [0.72-2.10] |
| Urine test | 1361 | 1.16 | [0.75-1.80] | 1361 | 1.28 | [0.80-2.06] |
| Blood test | 1361 | 1.2 | [0.81-1.78] | 1361 | 1.48 | [0.93-2.36] |
| Stool test | 1361 | 1.17 | [0.80-1.73] | 1361 | 1.27 | [0.83-1.95] |
| Syphilis test | 1361 | 0.7 | [0.43-1.14] | 1361 | 0.71 | [0.43-1.17] |
| HIV test | 1361 | 0.99 | [0.63-1.55] | 1361 | 1.2 | [0.69-2.07] |
| TT shot | 1361 | 1.24 | [0.80-1.91] | 1361 | 1.21 | [0.77-1.90] |
| IFA | 1361 | 1.16 | [0.69-1.96] | 1361 | 1.26 | [0.73-2.18] |
| Deworming | 1361 | 1.4 | [0.89-2.22] | 1356 | 1.53 | [0.96-2.45] |
| Women that received IFA during pregnancy | 1809 | 1.3 | [0.86-1.96] | 1809 | 1.49 | [0.96-2.31] |
| Women that received deworming during pregnancy | 1809 | 1.4 | [0.89-2.19] | 1800 | 1.56 | [0.99-2.45] |
| Pregnant women that sought care for: |  |  |  |  |  |  |
| Pregnancy complications | 918 | 2.15 | [1.39-3.32] | 918 | 2.2 | [1.41-3.43] |
| Delivery complications | 692 | 1.99 | [1.15-3.44] | 692 | 2.27 | [1.22-4.23] |
| Post-delivery complications | 556 | 3.49 | [1.89-6.45] | 556 | 3.89 | [1.95-7.77] |
| Women who delivered in a health facility | 1809 | 1.06 | [0.73-1.54] | 1809 | 1.07 | [0.66-1.75] |
| Among women delivering in a health facility: |  |  |  |  |  |  |
| C-section | 1103 | 0.79 | [0.40-1.56] | 1101 | 0.83 | [0.43-1.59] |
| Blood transfusion | 1103 | 1.63 | [0.33-8.07] | 702 | 1.38 | [0.33-5.74] |
| Uterotonic use | 1103 | 2.41 | [1.09-5.34] | 1101 | 2.67 | [1.13-6.31] |
| Mother checked after birth | 1103 | 1.19 | [0.75-1.89] | 1103 | 1.2 | [0.75-1.93] |
| Baby resuscitated with ambu bag# | 43 | 16.69 | [1.17-237.27] | 33 | - | - |
| Chlorohexidine applied to cord stump | 1083 | 0.73 | [0.28-1.93] | 1083 | 0.82 | [0.31-2.13] |
| Baby weighed at birth | 1103 | 1.33 | [0.76-2.34] | 1101 | 1.42 | [0.79-2.55] |
| Baby checked after birth | 1106 | 1 | [0.60-1.68] | 1106 | 1.01 | [0.60-1.69] |
| Skin to skin | 1106 | 0.83 | [0.46-1.51] | 1104 | 0.86 | [0.47-1.56] |
| Delayed bathing | 1106 | 1.66 | [0.89-3.09] | 1104 | 1.75 | [0.91-3.38] |
| Early initiation of breastfeeding | 1106 | 0.89 | [0.52-1.50] | 1104 | 0.97 | [0.55-1.70] |
| Women who received a c-section | 1809 | 0.83 | [0.43-1.57] | 1800 | 0.84 | [0.43-1.66] |
| Women who received a uterotonic | 1809 | 1.43 | [0.97-2.10] | 1809 | 1.63 | [1.07-2.48] |
| Newborns who had chlorohexidine applied to stump | 1779 | 0.98 | [0.42-2.28] | 1779 | 1.2 | [0.50-2.88] |
| Newborns receiving skin to skin | 1808 | 0.99 | [0.66-1.48] | 1808 | 1.04 | [0.65-1.65] |
| Newborns with delayed bathing | 1808 | 1.07 | [0.72-1.61] | 1808 | 1.14 | [0.74-1.75] |
| Newborns with early initiation of breastfeeding | 1808 | 0.88 | [0.63-1.24] | 1808 | 0.92 | [0.65-1.30] |
| Women who received a postnatal check within 48 hrs | 1809 | 1.28 | [0.91-1.81] | 1809 | 1.38 | [0.94-2.02] |
| Newborns who received a postnatal within first 48 hrs | 1779 | 1.24 | [0.83-1.85] | 1779 | 1.28 | [0.83-1.98] |
| Home visit (or sought care) within first week | 1779 | 1.37 | [0.84-2.24] | 1779 | 1.54 | [0.94-2.53] |
| BF counseling during PNC | 607 | 1.14 | [0.63-2.07] | 605 | 1.11 | [0.61-2.02] |
| Newborns who received BCG vaccine* | 1808 | 1.67 | [1.12-2.48] | 1808 | 2.22 | [1.35-3.67] |
|  | n | OR | 95% CI | n | AOR | 95% CI |
| Newborns who received polio vaccine* | 1808 | 1.8 | [1.28-2.54] | 1808 | 2.19 | [1.47-3.25] |
| Newborns exclusively breastfed* | 1761 | 0.94 | [0.62-1.41] | 1761 | 1.05 | [0.69-1.59] |
| Sought skilled care for NN illness | 641 | 1.65 | [1.00-2.72] | 638 | 1.83 | [1.07-3.11] |
| Women practicing family planning post-delivery* | 1809 | 1.74 | [1.17-2.58] | 1809 | 1.76 | [1.12-2.76] |
| Women who intend to practice family planning in next year* | 1492 | 1 | [0.72-1.38] | 1492 | 1.06 | [0.73-1.53] |

#among children in need of neonatal resuscitation based on maternal report of asphyxia at birth
\*at time of follow-up interview

**Table 3.**
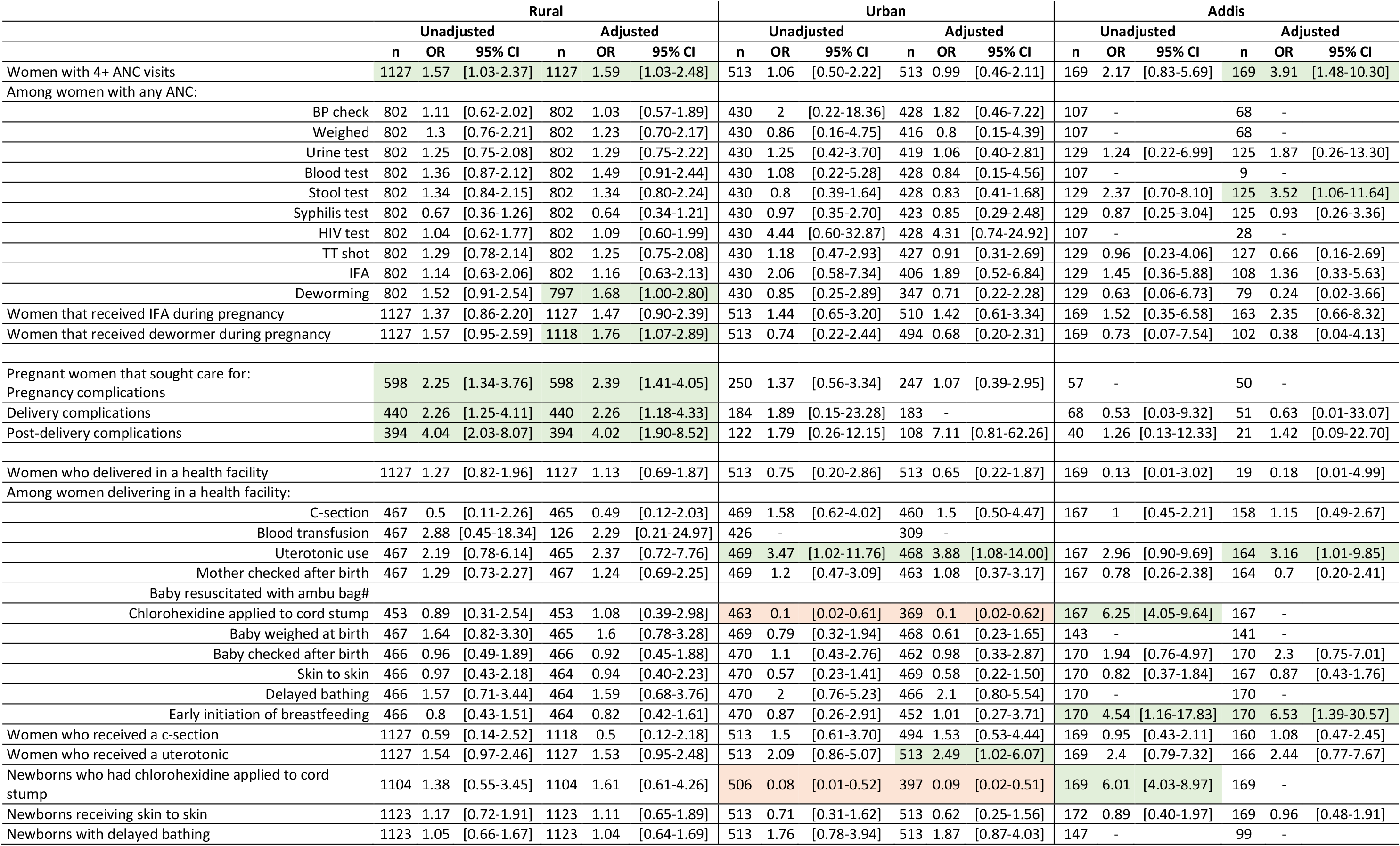
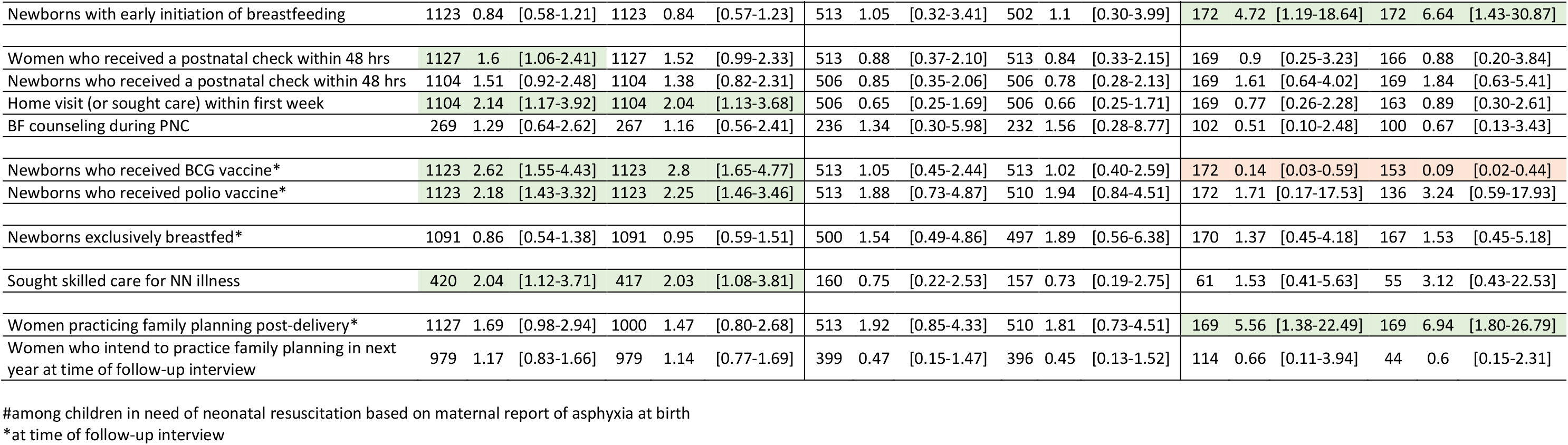
Odds of intervention receipt in COVID-19 impacted cohort versus unaffected reference cohort by urban (Addis and other urban areas) and rural areas.

|  | Rural |  |  |  |  |  | Urban |  |  |  |  |  | Addis |  |  |  |  |  |
| --- | --- | --- | --- | --- | --- | --- | --- | --- | --- | --- | --- | --- | --- | --- | --- | --- | --- | --- |
|  | Unadjusted |  |  | Adjusted |  |  | Unadjusted |  |  | Adjusted |  |  | Unadjusted |  |  | Adjusted |  |  |
|  | n | OR | 95% CI | n | OR | 95% CI | n | OR | 95% CI | n | OR | 95% CI | n | OR | 95% CI | n | OR | 95% CI |
| Women with 4+ ANC visits | 1127 | 1.57 | [1.03-2.37] | 1127 | 1.59 | [1.03-2.48] | 513 | 1.06 | [0.50-2.22] | 513 | 0.99 | [0.46-2.11] | 169 | 2.17 | [0.83-5.69] | 169 | 3.91 | [1.48-10.30] |
| Among women with any ANC: |  |  |  |  |  |  |  |  |  |  |  |  |  |  |  |  |  |  |
| BP check | 802 | 1.11 | [0.62-2.02] | 802 | 1.03 | [0.57-1.89] | 430 | 2 | [0.22-18.36] | 428 | 1.82 | [0.46-7.22] | 107 | - |  | 68 | - |  |
| Weighed | 802 | 1.3 | [0.76-2.21] | 802 | 1.23 | [0.70-2.17] | 430 | 0.86 | [0.16-4.75] | 416 | 0.8 | [0.15-4.39] | 107 | - |  | 68 | - |  |
| Urine test | 802 | 1.25 | [0.75-2.08] | 802 | 1.29 | [0.75-2.22] | 430 | 1.25 | [0.42-3.70] | 419 | 1.06 | [0.40-2.81] | 129 | 1.24 | [0.22-6.99] | 125 | 1.87 | [0.26-13.30] |
| Blood test | 802 | 1.36 | [0.87-2.12] | 802 | 1.49 | [0.91-2.44] | 430 | 1.08 | [0.22-5.28] | 428 | 0.84 | [0.15-4.56] | 107 | - |  | 9 | - |  |
| Stool test | 802 | 1.34 | [0.84-2.15] | 802 | 1.34 | [0.80-2.24] | 430 | 0.8 | [0.39-1.64] | 428 | 0.83 | [0.41-1.68] | 129 | 2.37 | [0.70-8.10] | 125 | 3.52 | [1.06-11.64] |
| Syphilis test | 802 | 0.67 | [0.36-1.26] | 802 | 0.64 | [0.34-1.21] | 430 | 0.97 | [0.35-2.70] | 423 | 0.85 | [0.29-2.48] | 129 | 0.87 | [0.25-3.04] | 125 | 0.93 | [0.26-3.36] |
| HIV test | 802 | 1.04 | [0.62-1.77] | 802 | 1.09 | [0.60-1.99] | 430 | 4.44 | [0.60-32.87] | 428 | 4.31 | [0.74-24.92] | 107 | - |  | 28 | - |  |
| TT shot | 802 | 1.29 | [0.78-2.14] | 802 | 1.25 | [0.75-2.08] | 430 | 1.18 | [0.47-2.93] | 427 | 0.91 | [0.31-2.69] | 129 | 0.96 | [0.23-4.06] | 127 | 0.66 | [0.16-2.69] |
| IFA | 802 | 1.14 | [0.63-2.06] | 802 | 1.16 | [0.63-2.13] | 430 | 2.06 | [0.58-7.34] | 406 | 1.89 | [0.52-6.84] | 129 | 1.45 | [0.36-5.88] | 108 | 1.36 | [0.33-5.63] |
| Deworming | 802 | 1.52 | [0.91-2.54] | 797 | 1.68 | [1.00-2.80] | 430 | 0.85 | [0.25-2.89] | 347 | 0.71 | [0.22-2.28] | 129 | 0.63 | [0.06-6.73] | 79 | 0.24 | [0.02-3.66] |
| Women that received IFA during pregnancy | 1127 | 1.37 | [0.86-2.20] | 1127 | 1.47 | [0.90-2.39] | 513 | 1.44 | [0.65-3.20] | 510 | 1.42 | [0.61-3.34] | 169 | 1.52 | [0.35-6.58] | 163 | 2.35 | [0.66-8.32] |
| Women that received dewormer during pregnancy | 1127 | 1.57 | [0.95-2.59] | 1118 | 1.76 | [1.07-2.89] | 513 | 0.74 | [0.22-2.44] | 494 | 0.68 | [0.20-2.31] | 169 | 0.73 | [0.07-7.54] | 102 | 0.38 | [0.04-4.13] |
| Pregnant women that sought care for: |  |  |  |  |  |  |  |  |  |  |  |  |  |  |  |  |  |  |
| Pregnancy complications | 598 | 2.25 | [1.34-3.76] | 598 | 2.39 | [1.41-4.05] | 250 | 1.37 | [0.56-3.34] | 247 | 1.07 | [0.39-2.95] | 57 | - |  | 50 | - |  |
| Delivery complications | 440 | 2.26 | [1.25-4.11] | 440 | 2.26 | [1.18-4.33] | 184 | 1.89 | [0.15-23.28] | 183 | - |  | 68 | 0.53 | [0.03-9.32] | 51 | 0.63 | [0.01-33.07] |
| Post-delivery complications | 394 | 4.04 | [2.03-8.07] | 394 | 4.02 | [1.90-8.52] | 122 | 1.79 | [0.26-12.15] | 108 | 7.11 | [0.81-62.26] | 40 | 1.26 | [0.13-12.33] | 21 | 1.42 | [0.09-22.70] |
| Women who delivered in a health facility | 1127 | 1.27 | [0.82-1.96] | 1127 | 1.13 | [0.69-1.87] | 513 | 0.75 | [0.20-2.86] | 513 | 0.65 | [0.22-1.87] | 169 | 0.13 | [0.01-3.02] | 19 | 0.18 | [0.01-4.99] |
| Among women delivering in a health facility: |  |  |  |  |  |  |  |  |  |  |  |  |  |  |  |  |  |  |
| C-section | 467 | 0.5 | [0.11-2.26] | 465 | 0.49 | [0.12-2.03] | 469 | 1.58 | [0.62-4.02] | 460 | 1.5 | [0.50-4.47] | 167 | 1 | [0.45-2.21] | 158 | 1.15 | [0.49-2.67] |
| Blood transfusion | 467 | 2.88 | [0.45-18.34] | 126 | 2.29 | [0.21-24.97] | 426 | - |  | 309 | - |  |  |  |  |  |  |  |
| Uterotonic use | 467 | 2.19 | [0.78-6.14] | 465 | 2.37 | [0.72-7.76] | 469 | 3.47 | [1.02-11.76] | 468 | 3.88 | [1.08-14.00] | 167 | 2.96 | [0.90-9.69] | 164 | 3.16 | [1.01-9.85] |
| Mother checked after birth | 467 | 1.29 | [0.73-2.27] | 467 | 1.24 | [0.69-2.25] | 469 | 1.2 | [0.47-3.09] | 463 | 1.08 | [0.37-3.17] | 167 | 0.78 | [0.26-2.38] | 164 | 0.7 | [0.20-2.41] |
| Baby resuscitated with ambu bag# |  |  |  |  |  |  |  |  |  |  |  |  |  |  |  |  |  |  |
| Chlorohexidine applied to cord stump | 453 | 0.89 | [0.31-2.54] | 453 | 1.08 | [0.39-2.98] | 463 | 0.1 | [0.02-0.61] | 369 | 0.1 | [0.02-0.62] | 167 | 6.25 | [4.05-9.64] | 167 | - |  |
| Baby weighed at birth | 467 | 1.64 | [0.82-3.30] | 465 | 1.6 | [0.78-3.28] | 469 | 0.79 | [0.32-1.94] | 468 | 0.61 | [0.23-1.65] | 143 | - |  | 141 | - |  |
| Baby checked after birth | 466 | 0.96 | [0.49-1.89] | 466 | 0.92 | [0.45-1.88] | 470 | 1.1 | [0.43-2.76] | 462 | 0.98 | [0.33-2.87] | 170 | 1.94 | [0.76-4.97] | 170 | 2.3 | [0.75-7.01] |
| Skin to skin | 466 | 0.97 | [0.43-2.18] | 464 | 0.94 | [0.40-2.23] | 470 | 0.57 | [0.23-1.41] | 469 | 0.58 | [0.22-1.50] | 170 | 0.82 | [0.37-1.84] | 167 | 0.87 | [0.43-1.76] |
| Delayed bathing | 466 | 1.57 | [0.71-3.44] | 464 | 1.59 | [0.68-3.76] | 470 | 2 | [0.76-5.23] | 466 | 2.1 | [0.80-5.54] | 170 | - |  | 170 | - |  |
| Early initiation of breastfeeding | 466 | 0.8 | [0.43-1.51] | 464 | 0.82 | [0.42-1.61] | 470 | 0.87 | [0.26-2.91] | 452 | 1.01 | [0.27-3.71] | 170 | 4.54 | [1.16-17.83] | 170 | 6.53 | [1.39-30.57] |
| Women who received a c-section | 1127 | 0.59 | [0.14-2.52] | 1118 | 0.5 | [0.12-2.18] | 513 | 1.5 | [0.61-3.70] | 494 | 1.53 | [0.53-4.44] | 169 | 0.95 | [0.43-2.11] | 160 | 1.08 | [0.47-2.45] |
| Women who received a uterotonic | 1127 | 1.54 | [0.97-2.46] | 1127 | 1.53 | [0.95-2.48] | 513 | 2.09 | [0.86-5.07] | 513 | 2.49 | [1.02-6.07] | 169 | 2.4 | [0.79-7.32] | 166 | 2.44 | [0.77-7.67] |
| Newborns who had chlorohexidine applied to cord stump | 1104 | 1.38 | [0.55-3.45] | 1104 | 1.61 | [0.61-4.26] | 506 | 0.08 | [0.01-0.52] | 397 | 0.09 | [0.02-0.51] | 169 | 6.01 | [4.03-8.97] | 169 | - |  |
| Newborns receiving skin to skin | 1123 | 1.17 | [0.72-1.91] | 1123 | 1.11 | [0.65-1.89] | 513 | 0.71 | [0.31-1.62] | 513 | 0.62 | [0.25-1.56] | 172 | 0.89 | [0.40-1.97] | 169 | 0.96 | [0.48-1.91] |
| Newborns with delayed bathing | 1123 | 1.05 | [0.66-1.67] | 1123 | 1.04 | [0.64-1.69] | 513 | 1.76 | [0.78-3.94] | 513 | 1.87 | [0.87-4.03] | 147 | - |  | 99 | - |  |
|  | Unadjusted |  |  | Adjusted |  |  | Unadjusted |  |  | Adjusted |  |  | Unadjusted |  |  | Adjusted |  |  |
|  | n | OR | 95% CI | n | OR | 95% CI | n | OR | 95% CI | n | OR | 95% CI | n | OR | 95% CI | n | OR | 95% CI |
| Newborns with early initiation of breastfeeding | 1123 | 0.84 | [0.58-1.21] | 1123 | 0.84 | [0.57-1.23] | 513 | 1.05 | [0.32-3.41] | 502 | 1.1 | [0.30-3.99] | 172 | 4.72 | [1.19-18.64] | 172 | 6.64 | [1.43-30.87] |
| Women who received a postnatal check within 48 hrs | 1127 | 1.6 | [1.06-2.41] | 1127 | 1.52 | [0.99-2.33] | 513 | 0.88 | [0.37-2.10] | 513 | 0.84 | [0.33-2.15] | 169 | 0.9 | [0.25-3.23] | 166 | 0.88 | [0.20-3.84] |
| Newborns who received a postnatal check within 48 hrs | 1104 | 1.51 | [0.92-2.48] | 1104 | 1.38 | [0.82-2.31] | 506 | 0.85 | [0.35-2.06] | 506 | 0.78 | [0.28-2.13] | 169 | 1.61 | [0.64-4.02] | 169 | 1.84 | [0.63-5.41] |
| Home visit (or sought care) within first week | 1104 | 2.14 | [1.17-3.92] | 1104 | 2.04 | [1.13-3.68] | 506 | 0.65 | [0.25-1.69] | 506 | 0.66 | [0.25-1.71] | 169 | 0.77 | [0.26-2.28] | 163 | 0.89 | [0.30-2.61] |
| BF counseling during PNC | 269 | 1.29 | [0.64-2.62] | 267 | 1.16 | [0.56-2.41] | 236 | 1.34 | [0.30-5.98] | 232 | 1.56 | [0.28-8.77] | 102 | 0.51 | [0.10-2.48] | 100 | 0.67 | [0.13-3.43] |
| Newborns who received BCG vaccine* | 1123 | 2.62 | [1.55-4.43] | 1123 | 2.8 | [1.65-4.77] | 513 | 1.05 | [0.45-2.44] | 513 | 1.02 | [0.40-2.59] | 172 | 0.14 | [0.03-0.59] | 153 | 0.09 | [0.02-0.44] |
| Newborns who received polio vaccine* | 1123 | 2.18 | [1.43-3.32] | 1123 | 2.25 | [1.46-3.46] | 513 | 1.88 | [0.73-4.87] | 510 | 1.94 | [0.84-4.51] | 172 | 1.71 | [0.17-17.53] | 136 | 3.24 | [0.59-17.93] |
| Newborns exclusively breastfed* | 1091 | 0.86 | [0.54-1.38] | 1091 | 0.95 | [0.59-1.51] | 500 | 1.54 | [0.49-4.86] | 497 | 1.89 | [0.56-6.38] | 170 | 1.37 | [0.45-4.18] | 167 | 1.53 | [0.45-5.18] |
| Sought skilled care for NN illness | 420 | 2.04 | [1.12-3.71] | 417 | 2.03 | [1.08-3.81] | 160 | 0.75 | [0.22-2.53] | 157 | 0.73 | [0.19-2.75] | 61 | 1.53 | [0.41-5.63] | 55 | 3.12 | [0.43-22.53] |
| Women practicing family planning post-delivery* | 1127 | 1.69 | [0.98-2.94] | 1000 | 1.47 | [0.80-2.68] | 513 | 1.92 | [0.85-4.33] | 510 | 1.81 | [0.73-4.51] | 169 | 5.56 | [1.38-22.49] | 169 | 6.94 | [1.80-26.79] |
| Women who intend to practice family planning in next year at time of follow-up interview | 979 | 1.17 | [0.83-1.66] | 979 | 1.14 | [0.77-1.69] | 399 | 0.47 | [0.15-1.47] | 396 | 0.45 | [0.13-1.52] | 114 | 0.66 | [0.11-3.94] | 44 | 0.6 | [0.15-2.31] |

#among children in need of neonatal resuscitation based on maternal report of asphyxia at birth
\*at time of follow-up interview

### Antenatal Care

At a national level, the adjusted odds ratio (AOR 1.53; 95% CI: 1.05-2.23) suggests a higher proportion of women in the COVID affected cohort received four or more ANC visits. Nationally, 39.4% (95% CI: 34.6- 44.3) of women in the reference cohort had at least four ANC visits compared to 46.7% (95% CI: 38.1- 55.5) in the COVID-19 affected cohort. The odds of more than four visits were also significantly higher among COVID-affected women within the rural population (AOR 1.59; 95% CI 1.03 2.48) and in Addis Ababa (AOR 3.91; 95% CI 1.48-10.30).

Despite a greater proportion receiving the recommended four or more visits, there was little difference in the content of ANC services they reported receiving. Nationally, there was no difference in the content of care. In rural areas, a greater proportion of women in the COVID-19 affected cohort reported receiving a deworming during the pregnancy (AOR 1.76; 95% CI 1.07-2.89), and in Addis Ababa, a greater proportion of women in the COVID-19 affected cohort who accessed ANC reported receiving a stool test (AOR 3.52; 95% CI 1.06-11.64).

### Care-seeking for complications

A consistently greater proportion of women in the COVID-19 affected cohort reported seeking care for complications during pregnancy, delivery, and post-delivery. Both the adjusted and unadjusted models showed greater odds of care-seeking for pregnancy complications (AOR 2.20; 95% CI 1.41-3.43), complications during delivery (AOR 2.27; 95% CI 1.22-4.23), and post-delivery complications (AOR 3.89; 95% CI 1.95-7.77). The association was driven by increased care-seeking for complications in rural areas, where the adjusted and unadjusted odds of care-seeking for pregnancy complications (AOR 2.39; 95% CI 1.41-4.05), complications during delivery (AOR 2.26; 95% CI 1.18-4.33), and post-delivery complications (AOR 4.02; 95% CI 1.90-8.52) were also higher in the COVID-19 affected cohort. There was no significant difference in care-seeking for complications in the urban population.

### Labor & Delivery and immediate newborn care

There was no difference in the overall facility delivery rate at the national level or within any of the stratified populations. Nationally, 55.5% (95% CI 44.9-65.6) of pregnant women delivered at a health facility in May 2020 or later compared to 54% (95% CI 41.2-52.2) of women who delivered prior to February 2020. Among women delivering at a health facility, there was limited variation in care content between the two cohorts. Nationally, among women who gave birth at a facility, the odds of uterotonic receipt after delivery was higher in the COVID-19 affected cohort (AOR 2.67, 95% CI 1.13-6.31). This association was observed in Addis Ababa (AOR 3.16, 95% CI 1.01-9.85) and other urban areas (AOR 3.88, 95% CI 1.08-14.00), but not in rural areas. The odds of early breastfeeding initiation in Addis Ababa was also higher in the COVID-19 affected cohort (AOR 6.64; 95% CI 1.43-30.87) than in the unaffected cohort.

In urban areas, not including Addis Ababa, the odds of chlorhexidine application to a newborn’s cord stump were significantly lower in the COVID-19 affected cohort among both facility births (AOR 0.01; 95% CI 0.02-0.62) and the total population (AOR 0.09; 0.02-0.51). Chlorohexidine use was low (<10%) in urban areas in the COVID-19 unaffected cohort, and the drop translates to 0.9% (95% CI 0.2-4.3%) coverage among births in the urban population in May 2020 or later.

### Postnatal Care (Routine PNC, Immunization, Sick Newborn Care, and Breastfeeding)

At a national level, there was no difference by cohort in the proportion of women or newborns that received a postnatal check within 48 hours of delivery, either before release from a facility birth or through a visit with a health center or a health extension worker (HEW) following a community birth. In rural areas, the odds of receiving a home PNC visit (or check at a health facility) within the first week after birth was higher in the COVID-19 affected cohort (AOR 2.04; 95% CI 1.13-3.68) versus the reference cohort, doubling from 8% (95% CI 5.9-10.9%) to 15.8% (95% CI 9.1-25.9%) receiving a visit.

Nationally, a greater proportion of children in the COVID-19 affected cohort were reported to have received a BCG or polio vaccine by the time of follow-up interview. This was driven by an increase in the odds of immunization in the COVID-19 cohort in rural areas. The odds of both BCG (AOR 2.8; 95% CI 1.65-4.77) and polio (AOR 2.25; 95% CI 1.46-3.46) vaccination were higher in the rural COVID-19 affected cohort. However, in Addis Ababa, the odds of BCG vaccination were lower in the COVID-19 affected cohort compared to the reference cohort (AOR 0.09; 95% CI 0.02-0.44). In Addis Ababa, BCG vaccination coverage by six weeks of age dropped from 94.6% (95% CI 90.2-82.8%) to 71.1% (95% CI 40.9-89.8%) in the COVID-19 affected cohort.

In rural areas, the odds of care-seeking for sick newborns or young infants doubled in the COVID-19 affected cohort compared to the reference cohort (AOR 2.03; 95% CI 1.08-3.81). This represents an increase from 23.8% (17.4-31.7%) seeking care in the COVID-unaffected cohort to 38.9% (26.8-52.7%) seeking care in the COVID-19 affected cohort. There was no significant difference in care-seeking in the urban population, including Addis Ababa.

### Postpartum Family Planning

At a national level, the odds of a woman reporting she was using some form of family planning after her most recent birth were significantly greater in the COVID-19 affected cohort compared to the reference cohort. This was driven by a nearly seven-fold increase in reported postpartum family planning use in Addis Ababa among women who gave birth in May 2020 or later compared to women who gave birth before February 2020 (AOR 6.94; 95% CI 1.80-26.79). At six weeks after birth, 25.2% (18.9-32.7%) of women who gave birth prior to February 2020 reported using some form of postpartum family planning. This increased to 65.2% (36.1-86.1%) of women who delivered in May 2020 or later.

### Intervention Coverage Sensitivity Analysis

Supplemental Tables 3 and 4 show the odds ratio of intervention receipt in the COVID-19 affected and unaffected cohorts, using a more liberal definition of cohorts. Rather than excluding those respondents whose six-week follow-up interview was delayed, we included those interviews in this analysis treating births between August 2019 and February 2020 as our COVID-19 unaffected reference cohort and births in April 2020 or later as our COVID-19 affected cohort.

Expanding the cohort time periods did not notably alter the results for the reference period invariant indicators. The increased odds of uterotonic use in the COVID-19 affected cohort at a national level is non-significant in the sensitivity analysis, although it remains significantly associated in urban areas. The odds of chlorohexidine use are also not significantly lower in the COVID-19 affected cohort in urban areas in the sensitivity analysis.

Constraining the analysis of indicators with unrestricted reference periods to follow-up interviews that occurred between 5 to 10 weeks after birth, there is no longer a difference in vaccination coverage nationally. However, the adjusted odds of BCG vaccination in the COVID-19 affected cohort remained statistically greater in the rural population (AOR 2.06; 95% CI 1.07-3.95) and statistically lower in Addis Ababa (AOR: 0.05; 0.01-0.37) for the COVID-19 affected cohort. Contrary to our primary analysis, the sensitivity analysis found no difference in care-seeking for neonatal or young infant illness in any area and no national-level difference in postpartum family planning coverage. The sensitivity analysis also showed a reduced magnitude of the greater odds of postpartum family planning in the COVID-19 affected cohort in Addis Ababa (AOR 4.02; 95% CI 1.02-15.85).

### Mortality Outcomes

Beyond intervention receipt, we examined differences in maternally reported stillbirths and neonatal deaths in the two cohorts (Table 4). There was no significant difference in neonatal mortality. However, nationally, stillbirths were more common in the COVID-19 affected cohort (4.8%; 95% CI 2.6-8.6%) compared to the reference cohort (2.1%, 95% CI 1.1-4.2%). The odds of stillbirth were borderline higher in the COVID-19 affected cohort in the primary analysis, but the sensitivity analysis found the odds of stillbirth were 2.58 times higher (95% CI 1.04-6.43) among births in April 2020 or later compared to births prior to March 2020 (Supplemental Table 3).

**Table 4.** Differences in stillbirth and neonatal death rates in COVID-19 impacted cohort versus unaffected reference cohort by strata.

|  | Unadjusted |  |  | Adjusted |  |  | Births Aug 2019 - Jan 2020 |  |  | Births May 2020 + |  |  |
| --- | --- | --- | --- | --- | --- | --- | --- | --- | --- | --- | --- | --- |
|  | n | RR | 95% CI | n | ARR | 95% CI | n | Rate | 95% CI | n | Rate | 95% CI |
| **Stillbirths** |  |  |  |  |  |  |  |  |  |  |  |  |
| National | 785 | 2.24 | [0.93-5.39] | 785 | 2.71 | [0.92-7.94] | 523 | 0.021 | [0.011,0.042] | 262 | 0.048 | [0.026,0.086] |
| Rural | 536 | 2.03 | [0.79-5.18] | 536 | 2.19 | [0.72-6.70] | 348 | 0.024 | [0.012,0.050] | 188 | 0.049 | [0.025,0.094] |
| Urban | 186 | 5.19 | [0.30-88.65] | 186 | - |  | 137 | 0.009 | [0.001,0.086] | 49 | 0.049 | [0.011,0.197] |
| Addis Ababa | 63 | - |  | 63 | - |  | 147 | 0 | - | 25 | 0 | - |
| **Neonatal deaths** |  |  |  |  |  |  |  |  |  |  |  |  |
| National | 1808 | 0.99 | [0.38-2.56] | 1808 | 0.83 | [0.34-2.05] | 1,559 | 0.023 | [0.015,0.035] | 249 | 0.023 | [0.009,0.056] |
| Rural | 1123 | 0.78 | [0.24-2.55] | 1123 | 0.69 | [0.23-2.05] | 946 | 0.025 | [0.015,0.040] | 177 | 0.019 | [0.006,0.061] |
| Urban | 513 | 2.84 | [0.44-18.31] | 513 | - |  | 466 | 0.018 | [0.008,0.042] | 47 | 0.052 | [0.011,0.214] |
| Addis Ababa | 172 | - |  | 172 | - |  | 147 | 0.014 | [0.003,0.054] | 25 | 0 | - |

## Discussion

The availability of data on care from a representative sample of women who gave birth just before the COVID-19 pandemic and early in the pandemic offers unique insight into the impact of the early stages of the pandemic on peripartum care in Ethiopia. Unlike other data sources for monitoring the impact of the COVID-19 pandemic and response on maternal and child health, this study provides standard indicators of RMNH coverage from a representative sample of women who recently gave birth. If the pandemic, or pandemic response, disrupted access or use of health services, we expect it would be detectable in intervention coverage measures.

This dataset shows little evidence of COVID-19 disrupting RMNH services in Ethiopia in the initial few months of the pandemic. There were no significant reductions in the proportion of women seeking health services or the content of services they received for either preventative or curative interventions. In rural areas, the data suggest a greater proportion of women in the COVID-19 affected cohort sought care for pregnancy, delivery, and postpartum complications, as well as ANC, PNC, and care for sick newborns. Similar increases were not detectable in urban areas. The only significant reductions in coverage observed in the COVID-19 affected cohort were commodity-dependent interventions, specifically BCG vaccination in Addis Ababa and chlorohexidine use in other urban areas. The clearest evidence of a potential change in health behavior tied to the pandemic was the increased proportion of women in Addis Ababa who reported postpartum family planning in the COVID-19 affected cohort.

Despite the lack of evidence of a reduction in health services, the data suggest increased stillbirths in the COVID-19 affected cohort. In the primary analysis, the small sample size could not detect a significant difference in the odds of stillbirth in the two cohorts; however, with the increased sample in the sensitivity analysis, the association was significant. Multiple studies have shown an increase in stillbirth rates, particularly in LMICs, during the pandemic (18,19). In the absence of reductions in ANC and childbirth coverage, the origins of this increase in stillbirth are unclear. However, reductions in the quality and comprehensiveness of antenatal and delivery services may have occurred that are not captured through this analysis.

The national and regional governments of Ethiopia acted in a quick and coordinated manner to respond to COVID-19. Preparations for the COVID-19 response began in January (20). With the first case detected in Ethiopia in March 2020, compulsory quarantine, communications programs, school closures, public gathering bans, and city/region-specific restaurant and bar closures were initiated. In addition to restrictions to limit the spread of COVID-19, the government issued guidance to support the continued provision of essential health services (21) and began recruiting additional health workers and recalling retired health workers to absorb the anticipated strain on the health system (22). Supplemented by actions to limit impacts on transportation and the economy (20), these efforts addressed potential barriers to care and averted reductions in RMNH service coverage in the early stages of the pandemic.

Our analysis showed no difference in overall facility delivery rates; however, a previous analysis of this dataset demonstrated a reduction in hospital births in urban areas, with deliveries shifting to lower-level health facilities (23). This shift away from higher-level facilities, particularly those hospitals selected to handle COVID-19 patients, may have occurred with other interventions as well. Our analysis examining overall changes in coverage does not capture the shift in location of service receipt if it did not alter overall service coverage. This study is a natural experiment that capitalizes on the chance start of the COVID-19 pandemic during ongoing PMA Ethiopia data collection. As such, the study was not powered to capture differences between the COVID-19 affected and unaffected cohorts. Although sampled women gave birth either before or during the COVID-19 pandemic at random, the two cohorts had slightly different characteristics. Our estimates of the effect of COVID-19 on coverage accounted for differences in known covariates; however, there is potential for residual confounding. Our analysis used standard indicators on care-seeking and content of care. These indicators are based on maternal report of care and may be subject to recall errors or social desirability biases (24). However, data in both cohorts were collected using the same questions, and we anticipate reporting errors to be consistent between the two cohorts. While the pandemic may have also impacted the quality of services, we do not have robust data on the quality of care received. Given the timing of the pandemic relative to the survey, our analysis only captures a snapshot of the impact of COVID-19 during late pregnancy, childbirth, and early infancy. As ANC services should be accessed throughout the pregnancy, the impact of COVID-19 on cumulative ANC interventions would likely be attenuated by the undisrupted services in earlier trimesters. Finally, our primary factor for defining our two cohorts is time. Seasonality, other health programs, or secular changes in services may also have impacted access to care for women who delivered prior to February 2020. However, all the births occurred within a ten-month window, minimizing the potential impact of non-COVID-19 related changes in the health system.

## Conclusions

The government of Ethiopia’s prompt and well-constructed response to control the COVID-19 pandemic and ensure continuity of essential health services appears to have successfully averted negative impacts on maternal and neonatal care to a large extent. While this analysis cannot address the later effects of the pandemic, the evidence suggesting little disruption to RMNH services in the initial stages of the pandemic is promising. As progress continues to be made in the control of the pandemic, continued efforts are needed to ensure that essential health services are maintained and even strengthened to prevent indirect loss of life. Lessons learned from Ethiopia’s largely successful response will be important in preparing for future crises and planning for effective emergency response.

## Data Availability

All data underlying these analyses are publicly available at https://www.pmadata.org/data/available-datasets/request-access-dataset.

## Author contributions

ICMJE criteria for authorship read and met: EC, EQ, TR, AS, SS, and LZ. Conducted analysis: EC, EQ. Drafted paper: EC, EQ, and LZ. Agree with manuscript and conclusions: EC, EQ, TR, AS, SS, and LZ. All authors read, edited, and approved the manuscript.

## Competing Interests

We declare no competing interests in accordance with ICMJE Conflict of Interest Guidelines.

## Funding

This work was supported, in whole, by the Bill and Melinda Gates Foundation [INV 009466]. Under the grant conditions of the Foundation, a Creative Commons Attribution 4.0 Generic License has already been assigned to the Author Accepted Manuscript version that might arise from this submission.

## Ethics Approval

All procedures were approved by both the Addis Ababa University, College of Health Sciences [075/13/SPH] and Johns Hopkins Bloomberg School of Public Health [00009391] Institutional Review Boards.

## Patient and public involvement

Patients and/or the public were not involved in the design, or conduct, or reporting, or dissemination plans of this analysis.

## Patient consent for publication

Not required.

## Provenance and peer review

Not commissioned; externally peer reviewed.

## Notes

### Competing Interest Statement

The authors have declared no competing interest.

