## Supplemental for "Impact of the early stages of the COVID-19 pandemic on coverage of RMNH interventions in Ethiopia"

### Impact of the early stages of the COVID-19 pandemic on coverage of RMNH interventions in Ethiopia: Supplementary Tables and Figures

Emily Carter, Linnea Zimmerman, Ellie Qian, Tim Roberton, Assefa Seme, Solomon Shiferaw

**Supplemental Table 1. Health intervention coverage by cohort at national level**

| | Births Aug 2019 - Jan 2020 | | | Births May 2020 + | | |
|---|---|---|---|---|---|---|
| | n | Proportion | 95% CI | n | Proportion | 95% CI |
| Women with 4+ ANC visits | 1,550 | 0.394 | [0.346,0.443] | 259 | 0.467 | [0.381,0.555] |
| Among women with any ANC: | | | | | | |
| BP check | 1,163 | 0.833 | [0.788,0.871] | 198 | 0.842 | [0.747,0.905] |
| Weighed | 1,163 | 0.775 | [0.719,0.822] | 198 | 0.799 | [0.700,0.871] |
| Urine test | 1,163 | 0.54 | [0.480,0.600] | 198 | 0.578 | [0.463,0.685] |
| Blood test | 1,163 | 0.722 | [0.665,0.772] | 198 | 0.756 | [0.665,0.829] |
| Stool test | 1,163 | 0.271 | [0.236,0.310] | 198 | 0.304 | [0.227,0.394] |
| Syphilis test | 1,163 | 0.199 | [0.157,0.249] | 198 | 0.148 | [0.097,0.220] |
| HIV test | 1,163 | 0.607 | [0.545,0.666] | 198 | 0.604 | [0.480,0.716] |
| TT shot | 1,163 | 0.667 | [0.610,0.719] | 198 | 0.712 | [0.621,0.788] |
| IFA | 1,163 | 0.756 | [0.704,0.802] | 198 | 0.782 | [0.683,0.857] |
| Deworming | 1,163 | 0.168 | [0.133,0.210] | 198 | 0.221 | [0.151,0.312] |
| Women that received IFA during pregnancy | 1,550 | 0.627 | [0.575,0.675] | 259 | 0.686 | [0.580,0.775] |
| Women that received deworming during pregnancy | 1,550 | 0.137 | [0.109,0.171] | 259 | 0.182 | [0.122,0.263] |
| Pregnant women that sought care for: Pregnancy complications | 803 | 0.466 | [0.412,0.522] | 115 | 0.653 | [0.551,0.742] |
| Delivery complications | 607 | 0.607 | [0.527,0.682] | 85 | 0.754 | [0.632,0.846] |
| Post-delivery complications | 491 | 0.435 | [0.361,0.512] | 65 | 0.729 | [0.588,0.835] |
| Women who delivered in a health facility | 1,550 | 0.54 | [0.481,0.598] | 259 | 0.555 | [0.449,0.656] |
| Among women delivering in a health facility: | | | | | | |
| C-section | 956 | 0.107 | [0.086,0.133] | 147 | 0.087 | [0.048,0.154] |
| Blood transfusion | 956 | 0.01 | [0.005,0.020] | 147 | 0.016 | [0.004,0.061] |
| Uterotonic use | 956 | 0.735 | [0.685,0.780] | 147 | 0.87 | [0.759,0.934] |
| Mother checked after birth | 956 | 0.607 | [0.550,0.662] | 147 | 0.648 | [0.542,0.741] |
| Baby resuscitated with ambu bag# | 40 | 0.323 | [0.177,0.515] | 3 | 0.889 | [0.404,0.989] |
| Chlorohexidine applied to cord stump | 946 | 0.096 | [0.063,0.142] | 137 | 0.072 | [0.031,0.159] |
| Baby weighed at birth | 956 | 0.715 | [0.670,0.756] | 147 | 0.769 | [0.663,0.850] |
| Baby checked after birth | 967 | 0.522 | [0.465,0.578] | 139 | 0.523 | [0.394,0.648] |
| Skin to skin | 967 | 0.775 | [0.731,0.814] | 139 | 0.741 | [0.623,0.833] |
| Delayed bathing | 967 | 0.7 | [0.643,0.751] | 139 | 0.795 | [0.677,0.877] |
| Early initiation of breastfeeding | 967 | 0.822 | [0.792,0.848] | 139 | 0.803 | [0.706,0.874] |
| Women who received a c-section | 1,550 | 0.058 | [0.046,0.073] | 259 | 0.048 | [0.027,0.086] |
| Women who received a uterotonic | 1,550 | 0.411 | [0.362,0.461] | 259 | 0.498 | [0.391,0.606] |
| Newborns who had chlorohexidine applied to cord stump | 1,533 | 0.054 | [0.035,0.081] | 246 | 0.052 | [0.025,0.106] |
| Newborns receiving skin to skin | 1,559 | 0.461 | [0.413,0.510] | 249 | 0.458 | [0.355,0.566] |
| Newborns with delayed bathing | 1,559 | 0.554 | [0.497,0.609] | 249 | 0.571 | [0.461,0.675] |
| Newborns with early initiation of breastfeeding | 1,559 | 0.793 | [0.759,0.824] | 249 | 0.772 | [0.701,0.830] |
| Women who received a postnatal check within first 48 hrs | 1,550 | 0.364 | [0.318,0.414] | 259 | 0.424 | [0.332,0.520] |
| Newborns who received a postnatal check within first 48 hrs | 1,533 | 0.316 | [0.273,0.362] | 246 | 0.363 | [0.268,0.471] |

| | Births Aug 2019 - Jan 2020 | | | Births May 2020 + | | |
|---|---|---|---|---|---|---|
| | **n** | **Proportion** | **95% CI** | **n** | **Proportion** | **95% CI** |
| Home visit (or sought care) within first week | 1,533 | 0.13 | [0.108,0.156] | 246 | 0.17 | [0.110,0.253] |
| BF counseling during PNC | 518 | 0.442 | [0.379,0.508] | 89 | 0.475 | [0.331,0.623] |
| Newborns who received BCG vaccine* | 1,559 | 0.272 | [0.231,0.317] | 249 | 0.383 | [0.293,0.483] |
| Newborns who received polio vaccine* | 1,559 | 0.374 | [0.330,0.419] | 249 | 0.518 | [0.430,0.606] |
| Newborns exclusively breastfed* | 1,517 | 0.758 | [0.722,0.790] | 244 | 0.746 | [0.663,0.814] |
| Sought skilled care for NN illness | 546 | 0.286 | [0.229,0.351] | 95 | 0.399 | [0.292,0.516] |
| Women practicing family planning post-delivery* | 1,550 | 0.102 | [0.079,0.129] | 259 | 0.164 | [0.121,0.220] |
| Women who intend to practice family planning in next year at time of follow-up interview | 1,289 | 0.749 | [0.682,0.805] | 203 | 0.748 | [0.668,0.815] |

#among children in need of neonatal resuscitation based on maternal report of asphyxia at birth
*at time of follow-up interview

#### Supplemental Table 2. Health intervention coverage by cohort at national level by urban (Addis and other urban areas) and rural areas

| | Rural | | | | | | Urban | | | | | | Addis Ababa | | | | | |
|---|---|---|---|---|---|---|---|---|---|---|---|---|---|---|---|---|---|---|
| | Births Aug 2019 - Jan 2020 | | | Births May 2020 + | | | Births Aug 2019 - Jan 2020 | | | Births May 2020 + | | | Births Aug 2019 - Jan 2020 | | | Births May 2020 + | | |
| | n | Prop | 95% CI | n | Prop | 95% CI | n | Prop | 95% CI | n | Prop | 95% CI | n | Prop | 95% CI | n | Prop | 95% CI |
| Women with 4+ ANC visits | 942 | 0.317 | [0.265,0.373] | 185 | 0.42 | [0.326,0.522] | 464 | 0.64 | [0.526,0.740] | 49 | 0.652 | [0.451,0.811] | 144 | 0.697 | [0.469,0.857] | 25 | 0.834 | [0.571,0.950] |
| Among women with any ANC: | | | | | | | | | | | | | | | | | | |
| BP check | 666 | 0.796 | [0.738,0.844] | 136 | 0.813 | [0.702,0.890] | 390 | 0.938 | [0.865,0.972] | 40 | 0.968 | [0.753,0.997] | 107 | 0.98 | [0.924,0.995] | 22 | 1 | - |
| Weighed | 666 | 0.717 | [0.646,0.779] | 136 | 0.767 | [0.651,0.853] | 390 | 0.944 | [0.882,0.974] | 40 | 0.935 | [0.728,0.987] | 107 | 0.961 | [0.878,0.988] | 22 | 1 | - |
| Urine test | 666 | 0.481 | [0.408,0.555] | 136 | 0.537 | [0.405,0.664] | 390 | 0.698 | [0.579,0.796] | 40 | 0.743 | [0.493,0.896] | 107 | 0.825 | [0.664,0.919] | 22 | 0.854 | [0.583,0.961] |
| Blood test | 666 | 0.649 | [0.578,0.713] | 136 | 0.715 | [0.606,0.803] | 390 | 0.93 | [0.856,0.967] | 40 | 0.935 | [0.728,0.987] | 107 | 0.991 | [0.935,0.999] | 22 | 1 | - |
| Stool test | 666 | 0.235 | [0.194,0.282] | 136 | 0.292 | [0.204,0.399] | 390 | 0.411 | [0.338,0.488] | 40 | 0.357 | [0.202,0.550] | 107 | 0.206 | [0.102,0.374] | 22 | 0.381 | [0.225,0.567] |
| Syphilis test | 666 | 0.17 | [0.123,0.230] | 136 | 0.121 | [0.069,0.203] | 390 | 0.282 | [0.182,0.411] | 40 | 0.277 | [0.113,0.534] | 107 | 0.296 | [0.162,0.478] | 22 | 0.268 | [0.106,0.531] |
| HIV test | 666 | 0.513 | [0.441,0.586] | 136 | 0.524 | [0.384,0.661] | 390 | 0.867 | [0.759,0.931] | 40 | 0.967 | [0.806,0.995] | 107 | 0.99 | [0.928,0.998] | 22 | 1 | - |
| TT shot | 666 | 0.645 | [0.574,0.709] | 136 | 0.701 | [0.594,0.790] | 390 | 0.716 | [0.607,0.805] | 40 | 0.748 | [0.559,0.874] | 107 | 0.825 | [0.712,0.900] | 22 | 0.82 | [0.563,0.941] |
| IFA | 666 | 0.728 | [0.662,0.786] | 136 | 0.754 | [0.634,0.844] | 390 | 0.827 | [0.748,0.885] | 40 | 0.908 | [0.710,0.975] | 107 | 0.921 | [0.781,0.974] | 22 | 0.944 | [0.645,0.994] |
| Deworming | 666 | 0.174 | [0.131,0.228] | 136 | 0.242 | [0.162,0.347] | 390 | 0.165 | [0.106,0.247] | 40 | 0.143 | [0.038,0.411] | 107 | 0.067 | [0.024,0.173] | 22 | 0.043 | [0.007,0.235] |
| Women that received IFA during pregnancy | 942 | 0.575 | [0.512,0.635] | 185 | 0.65 | [0.526,0.757] | 464 | 0.775 | [0.695,0.839] | 49 | 0.832 | [0.682,0.920] | 144 | 0.927 | [0.821,0.973] | 25 | 0.951 | [0.680,0.994] |
| Women that received dewormer during pregnancy | 942 | 0.136 | [0.102,0.178] | 185 | 0.198 | [0.129,0.292] | 464 | 0.16 | [0.106,0.236] | 49 | 0.124 | [0.035,0.354] | 144 | 0.051 | [0.0]18,0.13] | 25 | 0.038 | [0.006,0.215] |
| Pregnant women that sought care for: Pregnancy complications | 519 | 0.464 | [0.397,0.533] | 79 | 0.661 | [0.539,0.764] | 227 | 0.442 | [0.371,0.515] | 23 | 0.52 | [0.319,0.715] | | | | | | |
| Delivery complications | 381 | 0.528 | [0.436,0.619] | 59 | 0.717 | [0.576,0.826] | 167 | 0.908 | [0.805,0.960] | 17 | 0.949 | [0.655,0.995] | 59 | 0.949 | [0.775,0.990] | 9 | 0.907 | [0.477,0.991] |
| Post-delivery complications | 346 | 0.372 | [0.291,0.460] | 48 | 0.705 | [0.540,0.830] | 112 | 0.745 | [0.593,0.854] | 10 | 0.839 | [0.432,0.973] | 33 | 0.841 | [0.661,0.935] | 7 | 0.87 | [0.467,0.981] |
| Women who delivered in a health facility | 942 | 0.427 | [0.359,0.498] | 185 | 0.486 | [0.366,0.607] | 464 | 0.899 | [0.830,0.942] | 49 | 0.87 | [0.572,0.971] | 144 | 0.994 | [0.957,0.999] | 25 | 0.958 | [0.715,0.995] |
| Among women delivering in a health facility: | | | | | | | | | | | | | | | | | | |
| C-section | 387 | 0.074 | [0.050,0.108] | 80 | 0.039 | [0.010,0.139] | 426 | 0.137 | [0.097,0.191] | 43 | 0.201 | [0.088,0.394] | 143 | 0.261 | [0.185,0.355] | 24 | 0.261 | [0.124,0.467] |
| Blood transfusion | 387 | 0.008 | [0.003,0.023] | 80 | 0.022 | [0.005,0.087] | 426 | 0.016 | [0.006,0.042] | 43 | 0 | - | 143 | 0 | - | 24 | 0 | - |
| Uterotonic use | 387 | 0.739 | [0.660,0.806] | 80 | 0.861 | [0.703,0.942] | 426 | 0.752 | [0.686,0.808] | 43 | 0.913 | [0.776,0.970] | 143 | 0.618 | [0.515,0.712] | 24 | 0.827 | [0.608,0.937] |
| Mother checked after birth | 387 | 0.568 | [0.492,0.641] | 80 | 0.629 | [0.493,0.747] | 426 | 0.64 | [0.523,0.742] | 43 | 0.681 | [0.477,0.834] | 143 | 0.801 | [0.671,0.888] | 24 | 0.759 | [0.542,0.893] |
| Baby resuscitated with ambu bag# | 17 | 0.329 | [0.148,0.581] | 1 | 1 | - | 15 | 0.282 | [0.050,0.746] | 1 | 1 | - | 8 | 0.404 | [0.140,0.738] | 1 | 0 | - |
| Chlorohexidine applied to cord stump | 381 | 0.104 | [0.063,0.167] | 72 | 0.094 | [0.038,0.212] | 422 | 0.099 | [0.041,0.217] | 41 | 0.01 | [0.002,0.050] | 143 | 0.006 | [0.001,0.051] | 24 | 0.038 | [0.005,0.233] |
| Baby weighed at birth | 387 | 0.635 | [0.573,0.692] | 80 | 0.74 | [0.596,0.846] | 426 | 0.834 | [0.746,0.895] | 43 | 0.798 | [0.651,0.893] | 143 | 0.863 | [0.778,0.919] | 24 | 1 | - |
| Baby checked after birth | 392 | 0.496 | [0.425,0.567] | 74 | 0.487 | [0.322,0.654] | 429 | 0.551 | [0.427,0.668] | 41 | 0.573 | [0.364,0.759] | 146 | 0.612 | [0.463,0.742] | 24 | 0.754 | [0.473,0.912] |
| Skin to skin | 392 | 0.765 | [0.697,0.821] | 74 | 0.758 | [0.594,0.870] | 429 | 0.798 | [0.734,0.849] | 41 | 0.691 | [0.506,0.830] | 146 | 0.756 | [0.663,0.829] | 24 | 0.718 | [0.544,0.845] |
| Delayed bathing | 392 | 0.688 | [0.604,0.761] | 74 | 0.775 | [0.617,0.881] | 429 | 0.664 | [0.574,0.743] | 41 | 0.797 | [0.588,0.916] | 146 | 0.974 | [0.933,0.990] | 24 | 1 | - |
| Early initiation of breastfeeding | 392 | 0.822 | [0.780,0.858] | 74 | 0.787 | [0.661,0.876] | 429 | 0.84 | [0.786,0.882] | 41 | 0.82 | [0.597,0.933] | 146 | 0.731 | [0.644,0.803] | 24 | 0.925 | [0.761,0.980] |
| Women who received a c-section | 942 | 0.032 | [0.021,0.047] | 185 | 0.019 | [0.005,0.069] | 464 | 0.123 | [0.085,0.175] | 49 | 0.174 | [0.077,0.350] | 144 | 0.26 | [0.183,0.354] | 25 | 0.25 | [0.118,0.454] |
| Women who received a uterotonic | 942 | 0.33 | [0.275,0.392] | 185 | 0.432 | [0.312,0.562] | 464 | 0.688 | [0.618,0.751] | 49 | 0.822 | [0.652,0.919] | 144 | 0.615 | [0.515,0.706] | 25 | 0.793 | [0.572,0.916] |
| Newborns who had chlorohexidine applied to cord stump | 930 | 0.044 | [0.026,0.074] | 174 | 0.06 | [0.028,0.126] | 459 | 0.099 | [0.042,0.214] | 47 | 0.009 | [0.002,0.043] | 144 | 0.006 | [0.001,0.051] | 25 | 0.036 | [0.005,0.226] |

| | Rural | | | | | | Urban | | | | | | Addis Ababa | | | | | |
|---|---|---|---|---|---|---|---|---|---|---|---|---|---|---|---|---|---|---|
| | Births Aug 2019 - Jan 2020 | | | Births May 2020 + | | | Births Aug 2019 - Jan 2020 | | | Births May 2020 + | | | Births Aug 2019 - Jan 2020 | | | Births May 2020 + | | |
| | **n** | **Prop** | **95% CI** | **n** | **Prop** | **95% CI** | **n** | **Prop** | **95% CI** | **n** | **Prop** | **95% CI** | **n** | **Prop** | **95% CI** | **n** | **Prop** | **95% CI** |
| Newborns receiving skin to skin | 946 | 0.373 | [0.318,0.433] | 177 | 0.411 | [0.293,0.540] | 466 | 0.748 | [0.685,0.801] | 47 | 0.677 | [0.502,0.813] | 147 | 0.752 | [0.663,0.823] | 25 | 0.73 | [0.557,0.853] |
| Newborns with delayed bathing | 946 | 0.509 | [0.439,0.578] | 177 | 0.521 | [0.395,0.645] | 466 | 0.648 | [0.561,0.725] | 47 | 0.763 | [0.583,0.882] | 147 | 0.971 | [0.933,0.990] | 25 | 1 | - |
| Newborns with early initiation of breastfeeding | 946 | 0.785 | [0.740,0.823] | 177 | 0.753 | [0.671,0.820] | 466 | 0.838 | [0.792,0.876] | 47 | 0.845 | [0.649,0.941] | 147 | 0.732 | [0.645,0.805] | 25 | 0.928 | [0.768,0.981] |
| Women who received a postnatal check within first 48 hrs | 942 | 0.276 | [0.225,0.334] | 185 | 0.379 | [0.277,0.492] | 464 | 0.627 | [0.519,0.724] | 49 | 0.597 | [0.388,0.775] | 144 | 0.816 | [0.689,0.899] | 25 | 0.8 | [0.581,0.920] |
| Newborns who received a postnatal check within first 48 hrs | 930 | 0.238 | [0.193,0.290] | 174 | 0.321 | [0.214,0.450] | 459 | 0.562 | [0.451,0.667] | 47 | 0.522 | [0.316,0.721] | 144 | 0.618 | [0.468,0.748] | 25 | 0.722 | [0.455,0.890] |
| Home visit (or sought care) within first week | 930 | 0.08 | [0.059,0.109] | 174 | 0.158 | [0.091,0.259] | 459 | 0.267 | [0.197,0.351] | 47 | 0.191 | [0.090,0.358] | 144 | 0.424 | [0.295,0.565] | 25 | 0.361 | [0.170,0.610] |
| BF counseling during PNC | 209 | 0.395 | [0.306,0.491] | 60 | 0.458 | [0.294,0.631] | 220 | 0.453 | [0.350,0.560] | 16 | 0.526 | [0.195,0.836] | 89 | 0.753 | [0.659,0.828] | 13 | 0.608 | [0.286,0.857] |
| Newborns who received BCG vaccine* | 946 | 0.162 | [0.121,0.214] | 177 | 0.337 | [0.235,0.456] | 466 | 0.569 | [0.452,0.679] | 47 | 0.581 | [0.393,0.748] | 147 | 0.946 | [0.902,0.971] | 25 | 0.711 | [0.409,0.898] |
| Newborns who received polio vaccine* | 946 | 0.281 | [0.234,0.333] | 177 | 0.459 | [0.358,0.563] | 466 | 0.626 | [0.504,0.733] | 47 | 0.759 | [0.547,0.891] | 147 | 0.944 | [0.897,0.970] | 25 | 0.967 | [0.773,0.996] |
| Newborns exclusively breastfed* | 917 | 0.763 | [0.718,0.803] | 174 | 0.736 | [0.639,0.814] | 455 | 0.773 | [0.708,0.828] | 45 | 0.84 | [0.628,0.942] | 145 | 0.571 | [0.469,0.668] | 25 | 0.646 | [0.432,0.814] |
| Sought skilled care for NN illness | 352 | 0.238 | [0.174,0.317] | 68 | 0.389 | [0.268,0.527] | 144 | 0.435 | [0.313,0.565] | 16 | 0.367 | [0.151,0.652] | 50 | 0.565 | [0.396,0.719] | 11 | 0.664 | [0.392,0.859] |
| Women practicing family planning post-delivery* | 942 | 0.076 | [0.052,0.111] | 185 | 0.123 | [0.080,0.184] | 464 | 0.174 | [0.125,0.238] | 49 | 0.288 | [0.155,0.473] | 144 | 0.252 | [0.189,0.327] | 25 | 0.652 | [0.361,0.861] |
| Women who intend to practice family planning in next year at time of follow-up interview | 817 | 0.719 | [0.637,0.788] | 162 | 0.749 | [0.663,0.819] | 367 | 0.847 | [0.764,0.905] | 32 | 0.722 | [0.389,0.914] | 105 | 0.927 | [0.828,0.971] | 9 | 0.893 | [0.658,0.973] |

#among children in need of neonatal resuscitation based on maternal report of asphyxia at birth
*at time of follow-up interview

**Supplemental Table 3. Odds of intervention receipt in COVID-19 impacted cohort (April 2020+ births) versus unaffected reference cohort (Aug 2019 – Feb 2020 births) at national level**

| | Unadjusted | | | Adjusted | | |
|---|---|---|---|---|---|---|
| | n | OR | 95% CI | n | AOR | 95% CI |
| Stillbirths | 1264 | 2.33 | [1.04-5.22] | 1262 | 2.58 | [1.04-6.43] |
| Neonatal deaths | 2292 | 1.81 | [0.94-3.47] | 2289 | 1.46 | [0.80-2.67] |
| Women with 4+ ANC visits | 2289 | 1.49 | [1.14-1.97] | 2286 | 1.72 | [1.26-2.35] |
| Among women with any ANC: | | | | | | |
| BP check | 1734 | 1.14 | [0.74-1.76] | 1734 | 1.13 | [0.72-1.78] |
| Weighed | 1734 | 0.97 | [0.64-1.45] | 1734 | 0.98 | [0.62-1.53] |
| Urine test | 1734 | 1.18 | [0.83-1.67] | 1734 | 1.3 | [0.91-1.86] |
| Blood test | 1734 | 1.09 | [0.82-1.46] | 1734 | 1.27 | [0.90-1.80] |
| Stool test | 1734 | 1.25 | [0.94-1.65] | 1734 | 1.33 | [0.98-1.80] |
| Syphilis test | 1734 | 0.94 | [0.65-1.36] | 1734 | 0.95 | [0.63-1.42] |
| HIV test | 1734 | 1.03 | [0.72-1.48] | 1734 | 1.2 | [0.80-1.80] |
| TT shot | 1734 | 1.42 | [0.98-2.06] | 1734 | 1.38 | [0.94-2.03] |
| IFA | 1734 | 1.12 | [0.74-1.69] | 1734 | 1.17 | [0.76-1.80] |
| Deworming | 1734 | 1.22 | [0.84-1.78] | 1734 | 1.26 | [0.87-1.82] |
| Women that received IFA during pregnancy | 2289 | 1.24 | [0.91-1.67] | 2286 | 1.35 | [0.97-1.88] |
| Women that received dewormer during pregnancy | 2289 | 1.24 | [0.86-1.79] | 2286 | 1.28 | [0.90-1.84] |
| Pregnant women that sought care for: Pregnancy complications | 1121 | 1.92 | [1.34-2.77] | 1121 | 1.94 | [1.34-2.83] |
| Delivery complications | 854 | 1.62 | [1.05-2.50] | 854 | 1.6 | [1.01-2.55] |
| Post-delivery complications | 685 | 3.2 | [1.98-5.16] | 685 | 3.08 | [1.89-5.03] |
| Women who delivered in a health facility | 2289 | 1.01 | [0.78-1.31] | 2286 | 1.04 | [0.75-1.45] |
| Among women delivering in a health facility: | | | | | | |
| C-section | 1406 | 0.83 | [0.51-1.36] | 1403 | 0.92 | [0.55-1.54] |
| Blood transfusion | 1406 | 1.7 | [0.49-5.88] | 1104 | 1.58 | [0.50-5.04] |
| Uterotonic use | 1406 | 1.53 | [0.90-2.59] | 1403 | 1.63 | [0.96-2.77] |
| Mother checked after birth | 1406 | 1.11 | [0.81-1.53] | 1406 | 1.19 | [0.87-1.62] |
| Baby resuscitated with ambu bag# | 59 | 1.85 | [0.34-10.12] | 54 | 0.35 | [0.01-13.66] |
| Chlorohexidine applied to cord stump | 1382 | 1.21 | [0.66-2.22] | 1382 | 1.22 | [0.66-2.25] |
| Baby weighed at birth | 1406 | 1.12 | [0.70-1.81] | 1403 | 1.19 | [0.73-1.92] |
| Baby checked after birth | 1413 | 1.15 | [0.82-1.61] | 1413 | 1.19 | [0.85-1.67] |
| Skin to skin | 1413 | 0.73 | [0.46-1.18] | 1410 | 0.74 | [0.47-1.19] |
| Delayed bathing | 1413 | 1.46 | [0.95-2.24] | 1410 | 1.57 | [1.00-2.46] |
| Early initiation of breastfeeding | 1413 | 0.87 | [0.60-1.25] | 1410 | 0.93 | [0.64-1.36] |
| Women who received a c-section | 2289 | 0.84 | [0.52-1.36] | 2274 | 0.91 | [0.54-1.53] |
| Women who received a uterotonic | 2289 | 1.18 | [0.91-1.53] | 2286 | 1.31 | [0.99-1.75] |
| Newborns who had chlorohexidine applied to cord stump | 2254 | 1.24 | [0.73-2.13] | 2251 | 1.29 | [0.78-2.13] |
| Newborns receiving skin to skin | 2293 | 0.92 | [0.68-1.25] | 2290 | 0.96 | [0.68-1.36] |
| Newborns with delayed bathing | 2293 | 1.13 | [0.86-1.49] | 2290 | 1.2 | [0.90-1.60] |
| Newborns with early initiation of breastfeeding | 2293 | 0.87 | [0.67-1.11] | 2290 | 0.9 | [0.70-1.16] |
| Women who received a postnatal check within first 48 hrs | 2289 | 1.19 | [0.94-1.51] | 2286 | 1.3 | [1.00-1.68] |
| Newborns who received a postnatal check within first 48 hrs | 2254 | 1.23 | [0.94-1.61] | 2251 | 1.32 | [0.98-1.77] |

| | Unadjusted | | | Adjusted | | |
|---|---|---|---|---|---|---|
| | n | OR | 95% CI | n | AOR | 95% CI |
| Home visit (or sought care) within first week | 2254 | 1.2 | [0.80-1.81] | 2251 | 1.33 | [0.89-2.01] |
| BF counseling during PNC | 785 | 1.52 | [0.97-2.40] | 783 | 1.49 | [0.96-2.32] |
| Newborns who received BCG vaccine* | 1498 | 1.12 | [0.69-1.83] | 1498 | 1.51 | [0.81-2.84] |
| Newborns who received polio vaccine* | 1498 | 0.9 | [0.57-1.42] | 1498 | 1.01 | [0.60-1.67] |
| Newborns exclusively breastfed* | 1460 | 0.89 | [0.55-1.44] | 1460 | 1.08 | [0.66-1.76] |
| Sought skilled care for NN illness* | 584 | 1.25 | [0.65-2.38] | 580 | 1.24 | [0.59-2.61] |
| Women practicing family planning post-delivery* | 1492 | 1.21 | [0.69-2.13] | 1492 | 1.17 | [0.64-2.15] |
| Women who intend to practice family planning in next year at time of follow-up interview | 1309 | 0.98 | [0.64-1.51] | 1309 | 1.04 | [0.63-1.73] |

#among children in need of neonatal resuscitation based on maternal report of asphyxia at birth
*at time of follow-up interview
+restricted to follow-up interviews between 5-10 weeks post birth

**Supplemental Table 4. Odds of intervention receipt in COVID-19 impacted cohort (April 2020+ births) versus unaffected reference cohort (Aug 2019 – Feb 2020 births) by urban (Addis and other urban areas) and rural areas**

| | Rural | | | | | | Urban | | | | | | Addis | | | | | |
|---|---|---|---|---|---|---|---|---|---|---|---|---|---|---|---|---|---|---|
| | Unadjusted | | | Adjusted | | | Unadjusted | | | Adjusted | | | Unadjusted | | | Adjusted | | |
| | n | OR | 95% CI | n | OR | 95% CI | n | OR | 95% CI | n | OR | 95% CI | n | OR | 95% CI | n | OR | 95% CI |
| Stillbirths | 815 | 2 | [0.83-4.81] | 813 | 2.05 | [0.76-5.52] | 338 | 5.5 | [0.52-58.64] | 338 | 6.14 | [6.14-6.14] | 111 | - | - | 111 | - | - |
| Neonatal deaths | 1408 | 1.91 | [0.93-3.94] | 1405 | 1.53 | [0.78-2.99] | 663 | 1.29 | [0.23-7.29] | 663 | 1.36 | [1.36-1.36] | 221 | 0 | - | 221 | 0 | - |
| Women with 4+ ANC visits | 1409 | 1.77 | [1.26-2.49] | 1406 | 1.86 | [1.29-2.70] | 661 | 1.04 | [0.58-1.85] | 661 | 0.99 | [0.56-1.77] | 219 | 1.92 | [0.62-5.95] | 219 | 2.71 | [0.90-8.11] |
| Among women with any ANC: | | | | | | | | | | | | | | | | | | |
| BP check | 1006 | 1.19 | [0.74-1.91] | 1006 | 1.08 | [0.67-1.76] | 559 | 1.57 | [0.58-4.24] | 555 | 1.94 | [0.75-4.98] | 130 | 1 | - | 84 | 1 | - |
| Weighed | 1006 | 1.07 | [0.68-1.67] | 1006 | 0.98 | [0.60-1.59] | 559 | 0.65 | [0.18-2.37] | 540 | 0.69 | [0.17-2.75] | 130 | 1 | - | 84 | 1 | - |
| Urine test | 1006 | 1.2 | [0.81-1.80] | 1006 | 1.23 | [0.82-1.85] | 559 | 1.62 | [0.63-4.15] | 544 | 1.72 | [0.71-4.20] | 169 | 1.27 | [0.23-7.06] | 165 | 1.85 | [0.30-11.29] |
| Blood test | 1006 | 1.2 | [0.86-1.67] | 1006 | 1.27 | [0.87-1.84] | 559 | 1.05 | [0.45-2.41] | 555 | 1.2 | [0.47-3.08] | 130 | 1 | - | 9 | 1 | - |
| Stool test | 1006 | 1.35 | [0.95-1.92] | 1006 | 1.32 | [0.92-1.90] | 559 | 1.01 | [0.56-1.85] | 555 | 1.05 | [0.60-1.84] | 169 | 2.6 | [1.03-6.52] | 165 | 3.55 | [1.18-10.68] |
| Syphilis test | 1006 | 0.96 | [0.58-1.58] | 1006 | 0.91 | [0.53-1.56] | 559 | 1 | [0.53-1.89] | 555 | 1.05 | [0.49-2.27] | 169 | 1.32 | [0.49-3.59] | 165 | 1.4 | [0.46-4.27] |
| HIV test | 1006 | 1.08 | [0.71-1.63] | 1006 | 1.09 | [0.70-1.71] | 559 | 3.2 | [1.09-9.38] | 555 | 4.84 | [1.69-13.90] | 169 | 0.38 | [0.02-6.23] | 111 | 0.48 | [0.09-2.47] |
| TT shot | 1006 | 1.5 | [0.96-2.35] | 1006 | 1.46 | [0.93-2.30] | 559 | 1.21 | [0.66-2.23] | 559 | 1 | [0.49-2.06] | 169 | 1.18 | [0.36-3.86] | 167 | 1.01 | [0.29-3.57] |
| IFA | 1006 | 1.08 | [0.67-1.74] | 1006 | 1.07 | [0.65-1.74] | 559 | 1.99 | [0.73-5.37] | 529 | 1.92 | [0.74-4.93] | 169 | 0.8 | [0.26-2.47] | 144 | 0.7 | [0.19-2.58] |
| Deworming | 1006 | 1.35 | [0.88-2.08] | 1006 | 1.44 | [0.94-2.20] | 559 | 0.72 | [0.35-1.49] | 533 | 0.7 | [0.36-1.39] | 169 | 0.43 | [0.04-5.10] | 97 | 0.21 | [0.02-2.62] |
| Women that received IFA during pregnancy | 1409 | 1.26 | [0.89-1.78] | 1406 | 1.28 | [0.88-1.86] | 661 | 1.75 | [0.86-3.55] | 657 | 1.75 | [0.86-3.56] | 219 | 0.92 | [0.30-2.84] | 209 | 0.99 | [0.30-3.22] |
| Women that received dewormer during pregnancy | 1409 | 1.42 | [0.94-2.14] | 1406 | 1.51 | [1.01-2.27] | 661 | 0.66 | [0.32-1.36] | 648 | 0.64 | [0.30-1.36] | 219 | 0.49 | [0.04-5.51] | 122 | 0.3 | [0.03-2.93] |
| Pregnant women that sought care for: | | | | | | | | | | | | | | | | | | |
| Pregnancy complications | 723 | 1.92 | [1.25-2.95] | 723 | 1.95 | [1.24-3.05] | 311 | 1.86 | [0.83-4.20] | 306 | 2.16 | [0.85-5.48] | 87 | 1.78 | [0.50-6.34] | 75 | 2.33 | [0.69-7.89] |
| Delivery complications | 534 | 1.87 | [1.17-3.00] | 534 | 1.61 | [0.99-2.62] | 235 | 0.85 | [0.12-6.03] | 233 | 0.65 | [0.10-4.15] | 85 | 0.82 | [0.05-13.03] | 58 | 0.68 | [0.02-30.29] |
| Post-delivery complications | 477 | 3.36 | [1.94-5.81] | 477 | 2.89 | [1.68-4.98] | 159 | 3.06 | [0.73-12.79] | 142 | 6.02 | [0.92-39.33] | 49 | 1.94 | [0.18-20.41] | 32 | 1.57 | [0.08-31.55] |
| Women who delivered in a health facility | 1409 | 1.18 | [0.86-1.61] | 1406 | 1.07 | [0.76-1.51] | 661 | 0.73 | [0.26-2.03] | 661 | 0.74 | [0.28-1.94] | 219 | 0.47 | [0.06-3.62] | 146 | 0.51 | [0.05-5.53] |
| Among women delivering in a health facility: | | | | | | | | | | | | | | | | | | |
| C-section | 583 | 0.66 | [0.23-1.90] | 580 | 0.73 | [0.24-2.19] | 610 | 1.23 | [0.62-2.43] | 598 | 1.28 | [0.60-2.74] | 213 | 0.99 | [0.51-1.95] | 213 | 1.07 | [0.50-2.27] |
| Blood transfusion | 583 | 2.1 | [0.37-11.99] | 583 | - | - | 610 | 0.81 | [0.06-10.24] | 610 | - | - | 213 | - | - | 213 | - | - |
| Uterotonic use | 583 | 1.22 | [0.60-2.46] | 580 | 1.23 | [0.58-2.59] | 610 | 2.74 | [1.34-5.61] | 608 | 2.84 | [1.35-6.00] | 213 | 3.11 | [1.17-8.25] | 209 | 3.36 | [1.18-9.56] |
| Mother checked after birth | 583 | 1.06 | [0.71-1.57] | 583 | 1.06 | [0.72-1.57] | 610 | 1.6 | [0.92-2.79] | 608 | 1.84 | [0.92-3.67] | 213 | 1.1 | [0.42-2.85] | 213 | 1.09 | [0.40-2.96] |
| Baby resuscitated with ambu bag# | 29 | - | - | 29 | - | - | 20 | - | - | 20 | - | - | 11 | - | - | 11 | - | - |
| Chlorohexidine applied to cord stump | 567 | 1.34 | [0.68-2.65] | 567 | 1.39 | [0.70-2.74] | 603 | 0.58 | [0.14-2.44] | 475 | 0.47 | [0.09-2.50] | 212 | 4.27 | [2.84-6.43] | 212 | - | - |
| Baby weighed at birth | 583 | 1.43 | [0.78-2.64] | 580 | 1.4 | [0.75-2.60] | 610 | 0.63 | [0.30-1.32] | 610 | 0.6 | [0.26-1.35] | 213 | 2.71 | [0.47-15.63] | 213 | 3.64 | [0.63-21.06] |
| Baby checked after birth | 585 | 1.06 | [0.68-1.64] | 585 | 1.04 | [0.66-1.64] | 613 | 1.5 | [0.82-2.74] | 611 | 1.66 | [0.77-3.57] | 215 | 1.82 | [0.63-5.30] | 215 | 2.1 | [0.64-6.95] |
| Skin to skin | 585 | 0.78 | [0.40-1.56] | 582 | 0.77 | [0.39-1.51] | 613 | 0.61 | [0.34-1.12] | 611 | 0.58 | [0.31-1.11] | 215 | 0.76 | [0.41-1.42] | 215 | 0.8 | [0.40-1.58] |
| Delayed bathing | 585 | 1.38 | [0.77-2.47] | 582 | 1.56 | [0.82-2.97] | 613 | 1.76 | [0.98-3.14] | 607 | 1.83 | [1.05-3.18] | 215 | 1.46 | [0.22-9.71] | 152 | 2.09 | [0.23-19.08] |
| Early initiation of breastfeeding | 585 | 0.81 | [0.52-1.27] | 582 | 0.88 | [0.54-1.45] | 613 | 0.81 | [0.37-1.78] | 591 | 0.86 | [0.38-1.95] | 215 | 2.8 | [1.24-6.33] | 215 | 3.44 | [1.28-9.25] |
| Women who received a c-section | 1409 | 0.74 | [0.26-2.07] | 1394 | 0.69 | [0.23-2.04] | 661 | 1.18 | [0.60-2.30] | 642 | 1.28 | [0.61-2.69] | 219 | 0.96 | [0.48-1.94] | 219 | 1.04 | [0.48-2.24] |
| Women who received a uterotonic | 1409 | 1.2 | [0.86-1.66] | 1406 | 1.17 | [0.83-1.64] | 661 | 1.9 | [1.06-3.40] | 661 | 2.09 | [1.19-3.65] | 219 | 2.55 | [1.01-6.46] | 215 | 2.73 | [1.01-7.36] |
| Newborns who had chlorohexidine applied to cord stump | 1383 | 1.54 | [0.87-2.72] | 1380 | 1.46 | [0.86-2.50] | 653 | 0.5 | [0.12-2.07] | 510 | 0.59 | [0.14-2.43] | 218 | 4.16 | [2.79-6.19] | 218 | - | - |
| Newborns receiving skin to skin | 1409 | 1.06 | [0.73-1.53] | 1406 | 1 | [0.66-1.51] | 663 | 0.74 | [0.41-1.34] | 663 | 0.71 | [0.39-1.31] | 221 | 0.77 | [0.41-1.45] | 221 | 0.83 | [0.42-1.65] |
| Newborns with delayed bathing | 1409 | 1.11 | [0.81-1.53] | 1406 | 1.13 | [0.81-1.57] | 663 | 1.73 | [0.98-3.04] | 663 | 1.81 | [1.05-3.10] | 221 | 1.5 | [0.22-10.28] | 158 | 2.09 | [0.19-22.84] |

| | Rural | | | | | | Urban | | | | | | Addis | | | | | |
|---|---|---|---|---|---|---|---|---|---|---|---|---|---|---|---|---|---|---|
| | Unadjusted | | | Adjusted | | | Unadjusted | | | Adjusted | | | Unadjusted | | | Adjusted | | |
| | n | OR | 95% CI | n | OR | 95% CI | n | OR | 95% CI | n | OR | 95% CI | n | OR | 95% CI | n | OR | 95% CI |
| Newborns with early initiation of breastfeeding | 1409 | 0.86 | [0.64-1.15] | 1406 | 0.88 | [0.66-1.17] | 663 | 0.8 | [0.39-1.64] | 650 | 0.81 | [0.39-1.67] | 221 | 2.29 | [1.11-4.74] | 221 | 2.78 | [1.12-6.87] |
| Women who received a postnatal check within first 48 hrs | 1409 | 1.38 | [1.04-1.84] | 1406 | 1.33 | [0.99-1.78] | 661 | 1.11 | [0.59-2.07] | 661 | 1.2 | [0.62-2.32] | 219 | 1.09 | [0.35-3.44] | 219 | 1.15 | [0.37-3.51] |
| Newborns who received a postnatal check within first 48 hrs | 1383 | 1.43 | [1.02-2.01] | 1380 | 1.35 | [0.95-1.93] | 653 | 1.09 | [0.57-2.06] | 653 | 1.12 | [0.54-2.33] | 218 | 1.51 | [0.56-4.04] | 218 | 1.74 | [0.59-5.15] |
| Home visit (or sought care) within first week | 1383 | 1.93 | [1.15-3.25] | 1380 | 1.88 | [1.12-3.15] | 653 | 0.5 | [0.22-1.13] | 653 | 0.54 | [0.23-1.29] | 218 | 0.74 | [0.28-1.90] | 218 | 0.84 | [0.33-2.16] |
| BF counseling during PNC | 353 | 1.74 | [1.01-3.00] | 351 | 1.62 | [0.94-2.81] | 301 | 1.33 | [0.46-3.88] | 296 | 1.19 | [0.40-3.50] | 131 | 0.68 | [0.23-1.99] | 131 | 0.9 | [0.29-2.87] |
| Newborns who received BCG vaccine*+ | 927 | 1.7 | [0.90-3.20] | 927 | 2.06 | [1.07-3.95] | 441 | 0.67 | [0.21-2.17] | 441 | 0.58 | [0.16-2.10] | 130 | 0.08 | [0.02-0.47] | 116 | 0.05 | [0.01-0.37] |
| Newborns who received polio vaccine*+ | 927 | 0.89 | [0.49-1.61] | 927 | 0.92 | [0.51-1.66] | 441 | 1.76 | [0.43-7.11] | 424 | 1.48 | [0.38-5.73] | 130 | 1.09 | [0.11-10.74] | 130 | - | - |
| Newborns exclusively breastfed*+ | 900 | 0.78 | [0.46-1.32] | 900 | 0.89 | [0.52-1.52] | 432 | 2.08 | [0.20-21.16] | 429 | 2.26 | [0.24-21.37] | 128 | 2 | [0.53-7.50] | 126 | 2.95 | [0.60-14.55] |
| Sought skilled care for NN illness+ | 385 | 1.27 | [0.57-2.83] | 381 | 1.09 | [0.43-2.75] | 148 | 1.05 | [0.19-5.86] | 143 | 0.89 | [0.22-3.61] | 51 | 2.65 | [0.53-13.21] | 46 | 4.23 | [0.54-33.14] |
| Women practicing family planning post-delivery*+ | 928 | 1.11 | [0.51-2.42] | 829 | 0.84 | [0.37-1.91] | 437 | 1.16 | [0.27-5.03] | 434 | 0.97 | [0.19-5.00] | 127 | 4.14 | [1.11-15.47] | 125 | 4.02 | [1.02-15.85] |
| Women who intend to practice family planning in next year at time of follow-up interview+ | 854 | 1.11 | [0.69-1.78] | 854 | 1.12 | [0.64-1.95] | 362 | 0.56 | [0.15-2.09] | 359 | 0.48 | [0.13-1.84] | 93 | 0.71 | [0.09-5.35] | 36 | 0.65 | [0.16-2.70] |

#among children in need of neonatal resuscitation based on maternal report of asphyxia at birth
*at time of follow-up interview
+restricted to follow-up interviews between 5-10 weeks post birth

**Supplemental Figure 1. Time between birth and follow-up interview by date of birth**

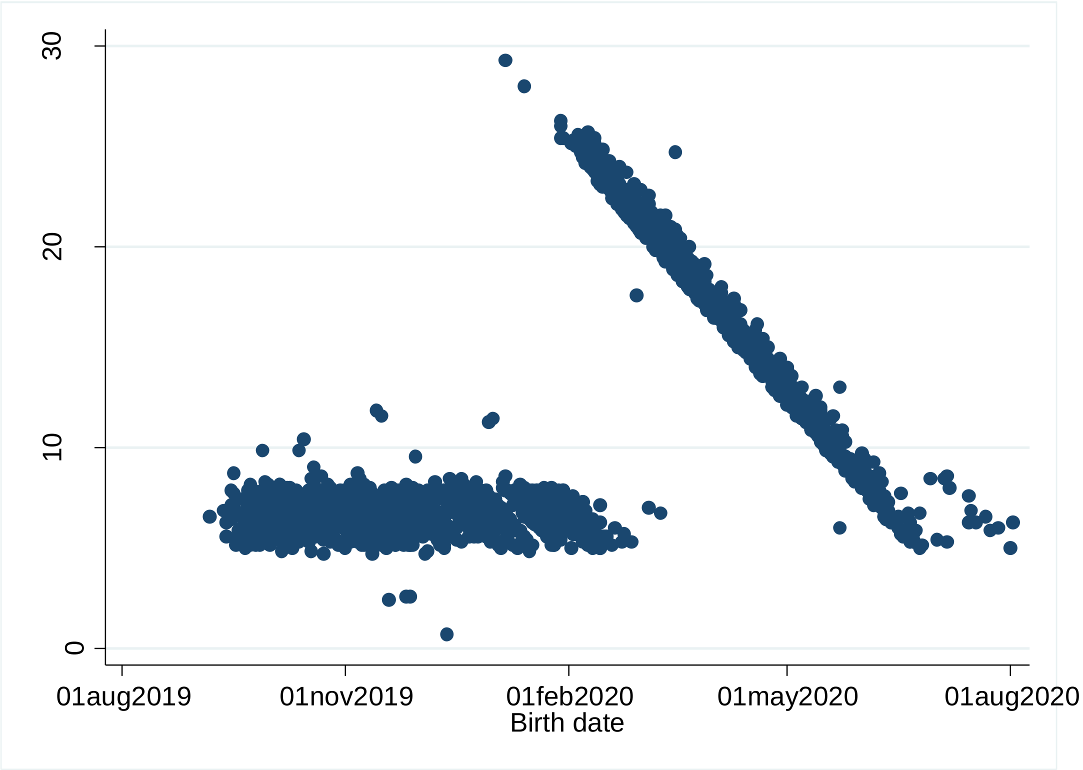

**Supplemental Figure 2. Number of births per month included in analysis**

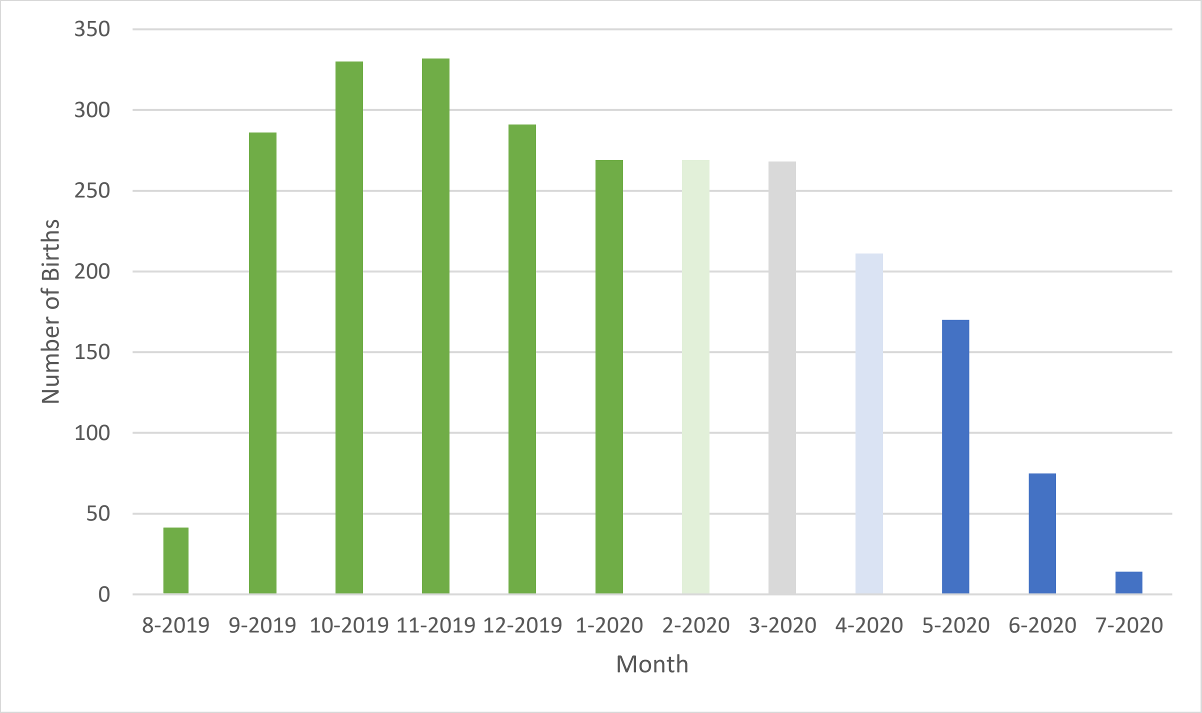

COVID-19 unaffected reference cohort in green. COVID-19 affected cohort in blue. Lighter shading indicates births only included in sensitivity analysis. March 2020 births (grey) excluded from all analyses.
